## Supplementary Material for "Different forms of superspreading lead to different outcomes: heterogeneity in infectiousness and contact behavior relevant for the case of SARS-CoV-2"

### S1 Population and parameters

#### S1.1 Population

We used a synthetic population of 11 million individuals, representing the total population of Belgium. The population is constructed once before all simulations are conducted. No births, deaths, immigration or emigration occur over the course of the simulations – making our population a closed one.

Household sizes and age constitutions were based on census data collected in Belgium in 2011 [1]. The geographic location of households is based on census data collected in 2001 [2]. Individuals in the population were assigned to schools and workplaces based on their geographic location, and school attendance and commuting data [2, 3],. Furthermore, each individual was assigned to two communities – to represent other contacts made during respectively the week and the weekend. These communities consist of, on average, 500 individuals, and are also based on geographic location.

#### S1.2 Social contact rates

Pre-pandemic contact rates were based on a social contact study conducted in Belgium in 2010 and 2011 [4]. We assumed households to be fully connected (i.e. the contact probability between two household members is always 0.999) [5].

Symptomatic individuals reduce their contacts at the workplace and at school by 90%, and have 75% less contacts in their community contact pools [6].

Contact reductions during lockdown periods, and periods of partial release were estimated based on social contact data collected for Belgium in the CoMix study [7]. To obtain contact rates during the lockdown period, we used results from surveys conducted from April to mid-May 2020, while we used results from surveys conducted from mid-May to August 2020 to obtain contact rates during the period of partial release.

Contact rates collected in the CoMix study were subdivided into different categories. We used contact rates from the 'work' category to estimate contact reductions in workplace contact pools, and contact rates from the 'transport', 'leisure', and 'other' categories to estimate contact reductions in the community social contact pools.

To estimate the contact reductions, we aggregated the contact rates during the lockdown period and the partial release phase over all ages, and compared these rates to the pre-pandemic contact rates as obtained from the social contact study conducted in 2010–2011 [4].

##### S1.3 Natural history of COVID-19

The natural history of COVID-19 was implemented in accordance with an earlier study conducted with STRIDE [8]. Following infection, individuals experience a latent period followed by either a pre-symptomatic and a symptomatic infectious period, or a fully asymptomatic infectious period.

The length of the incubation period (from the moment of infection to the start of the symptomatic/asymptomatic period) is drawn from a discretized log-normal distribution with logmean=1.43 and logsd=0.66 [9,10]. The length of the pre-symptomatic infectious period follows a Gamma distribution, with shape=20.52, rate=1.59 and shift=12.27, truncated at -1 and divided by the cumulative distribution function [9]. The total length of the infectious period is drawn from a normal distribution with mean=6 and sd=1 [9,11]. Finally, individuals that experience symptoms are symptomatic for 7 days, before they eventually recover [9].

Asymptomatic cases and individuals during the pre-symptomatic phase are assumed to be half as infectious as individuals experiencing symptoms [10].

The probability that an infected individual becomes symptomatic is age-dependent, and is calculated in the same way as for a previous study by Willem et al. [8,10,12].

Furthermore, we assumed that children are as susceptible as adults.

##### S1.4 Other relevant parameters

We chose February 17th 2020 as the start date for all simulations, and used holidays and school holidays as applicable in Belgium during the simulated period (2020–2021).

For further information about the model, and the manner in which it was adapted to model the transmission of SARS-CoV-2 we refer to previous work by Kuylen et al. (2017) and Willem et al. (2021) [8,13].

#### S2 Verification

##### S2.1 Distribution of secondary cases and $P_{80}$

We verified that the mean $R_0$ remained stable over the different scenarios. To estimate $R_0$, we looked at the mean of the secondary cases made by each index case in the simulations for the scenarios without interventions. In the violin plots in Supplementary Figure S1, the distribution and mean of the number of secondary cases caused per index case is shown. Variations in in the mean between the different scenarios are small, and are likely caused by stochasticity, as no upward or downward trend can be observed.

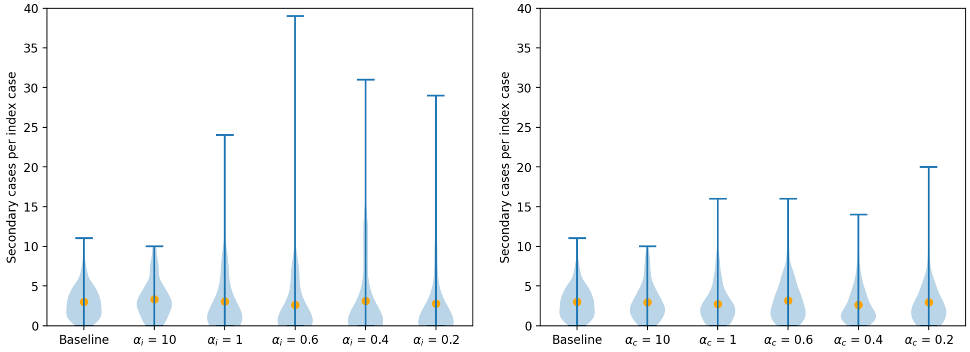


**(a)** Varying $\alpha_i$ for the Truncated Gamma distribution considered for the individual transmission probability.

**(b)** Varying $\alpha_c$ for the Gamma distribution considered the individual contact factor.

**Fig S1.** Violin plots for the number of secondary cases per index case for the different scenarios regarding infectiousness related heterogeneity (panel a) and contact-related heterogeneity (panel b). The orange dots represent the means of the simulated values.

We observe that, when increasing contact-related heterogeneity, the distribution of the estimated $R_0$ is less concentrated towards zero, and the maximum number of secondary cases is lower than it is when infectiousness-related heterogeneity is increased. As such, overdispersion seems to be less pronounced for the same values $\alpha_c$ compared to $\alpha_i$.

Once we were assured that the mean $R_0$ remained stable, we also verified that the variation in transmission changed accordingly when we increased either infectiousness-related or contact-related heterogeneity. To do this, we calculated $P_{80}$ (see Supplementary Figure S2), or the minimal proportion of infected individuals that is responsible for 80% of transmissions.

We observe that indeed, as we increase either infectiousness-related (Supplementary Figure S2a) or contact-related (Supplementary Figure S2b) heterogeneity, $P_{80}$ decreases, indicating that a smaller proportion of infected individuals is responsible for 80% of new cases. However, the effect is stronger when infectiousness-related heterogeneity is increased. When $\alpha_i$ is decreased from 10 to 0.2, $P_{80}$ decreases from 0.30 to 0.12, whereas $P_{80}$ decreases from 0.31 to 0.23 when $\alpha_c$ is decreased from 10 to 0.2. As such, we need a very high level of contact-related heterogeneity, or a certain level of infectiousness-related heterogeneity to obtain the range for $P_{80}$ (between 0.1 and 0.2) that was estimated for SARS-CoV-2 [14].

Finally, note that in the baseline case, $P_{80}$ is only 0.31, meaning that there already is a certain level of heterogeneity in transmission. This can be explained by the fact that in STRIDE, social mixing is never completely homogeneous, as contact rates are dependent on the ages of individuals and the locations in which contacts take place.

#### S2.2 Theoretical description

##### Theoretical description

We constructed a theoretical description to estimate the mean and variance of the number of secondary cases per infected individual. Let $i$ be a member of the (finite) population in Stride. Individual $i$ is assigned to different contact pools, such as a household or a workplace, in which they interact with other members of the population. We indicate the set of individuals that belong to a contact pool $k$, together with

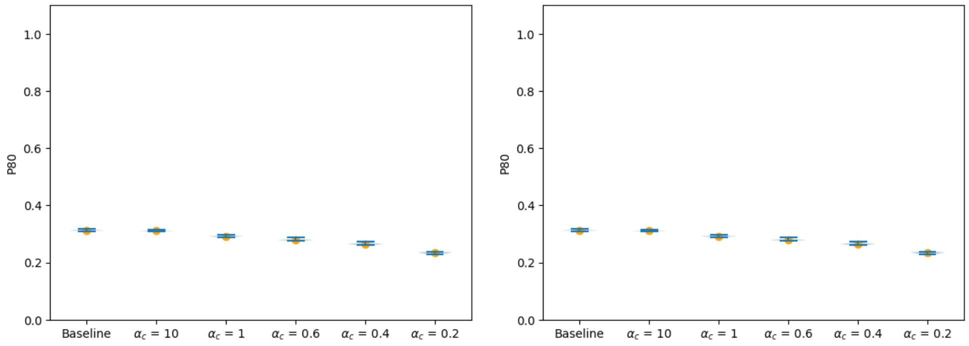


**(a)** Varying $\alpha_i$ for the Truncated Gamma distribution considered for the individual transmission probability.

**(b)** Varying $\alpha_c$ for the Gamma distribution considered for the individual contact factor.

**Fig S2.** Violin plots for $P_{80}$ over the different scenarios regarding infectiousness-related (panel a) and contact-related (panel b) heterogeneity. Runs which led to extinction (i.e. in which less than 20 cases occurred) were excluded.

individual $i$, as $L_k^i$. Then the collection of contact pools of which $i$ is a member, i.e. $\left\{L_k^i\right\}_k$, characterizes all individuals in the population that $i$ can contact.

In such a contact pool, individuals have a probability to contact other members. This probability depends on the age of the individuals, and the type of contact pool. Individuals who are members of the same contact pool have at most one contact per simulation day, according to a Bernouilli distribution. When a contact is realized between an infected and a susceptible individual, this contact can then result in an infection event with a defined probability – and as such this also follows a Bernouilli distribution.

We now consider $i$ an individual in a completely susceptible population, and $j$ and individual that shares a contact pool $\hat{k}$ with $i$, i.e. $j \in L_{\hat{k}}^i$. Assuming that $i$ starts their infectious period on day $t = 0$, and that they are infectious for $d$ days. In a single time-step, the probability that $i$ has an effective contact with $j$ can then be written as:

$$
\begin{aligned}
&\mathbb{P}(\text{“i has an effective contact with j”}) \\
=&\mathbb{P}(\text{“i has a contact with j”} \cap \text{“the contact between i and j is effective”}) \\
=&\mathbb{P}(\text{“the contact between i and j is effective”} \mid \text{“i has a contact with j”}) \\
&\times \mathbb{P}(\text{“i has a contact with j”}) \\
=&q_{i,j} c_{i,j}
\end{aligned}
$$

With $q_{i,j}$ the probability that a contact between $i$ and $j$ is effective, and $c_{i,j}$ is the probability that the individuals have a contact during the given time-step. According to this description, the number of effective contacts between $i$ and $j$, $X^{i,j}$, can be described by a Binomial distribution, $X^{i,j} \sim Bin(d, q_{i,j} c_{i,j})$, under the assumption that effective contact events are realized independently during the infectious period. When this is the case, we can write the probability that $i$ has at least one effective contact with $j$ as:

$$
\begin{aligned}
&\mathbb{P}(\text{“i has at least one effective contact with j”})\\
&= \mathbb{P}(X^{i,j} \geq 1)\\
&= 1 - \mathbb{P}(X^{i,j} < 1)\\
&= 1 - \mathbb{P}(X^{i,j} = 0)\\
&= 1 - (1 - q_{i,j}c_{i,j})^d
\end{aligned}
$$

Since we assume that the entire population, except for the index case $i$, is susceptible, the first effective contact between $i$ and $j$ will result in an infection event. Thus, the probability that $i$ infects $j$, $Y^{i,j}$, can be described by a Bernoulli random variable, $Y^{i,j} \sim Be(1 - (1 - q_{i,j}c_{i,j})^d)$. This description can be extended to account for all contacts that $i$ can possibly have in a given contact pool. The number of secondary cases that $i$ eventually makes in $\hat{k}$, $Y_{\hat{k}}$ is then given by:

$$Y_{\hat{k}} = \sum_{j \in L^i_{\hat{k}}, j \neq i} Y^{i,j}$$

Assuming that each $Y^{i,j}$ is independent from the others, $Y^{\hat{k}}$ follows a Poisson Binomial Distribution, with mean, $\mu$, and variance $\sigma^2$, given by:

$$
\begin{aligned}
\mu_{\hat{k}} &= \sum_{j \in L^i_{\hat{k}}, j \neq i} 1 - (1 - q_{i,j}c_{i,j})^d = |L^i_{\hat{k}}| - \sum_{j \in L^i_{\hat{k}}, j \neq i} (1 - q_{i,j}c_{i,j})^d\\
\sigma^2_{\hat{k}} &= \sum_{j \in L^i_{\hat{k}}, j \neq i} (1 - q_{i,j}c_{i,j})^d(1 - (1 - q_{i,j}c_{i,j})^d)
\end{aligned}
$$

Furthermore, the total number of secondary cases that $i$ can possibly generate within a susceptible population, $Y^i$, can be written as:

$$Y^i = \sum_{k \in K^i} \left( \sum_{j \in L^i_k, j \neq i} Y^{i,j} \right)$$

Wit $K^i$ the set of contact pools to which $i$ belongs. Same as above, $Y^i$, follows a Poisson Binomial distribution, with mean $\mu_i$ and variance $\sigma^2_i$, given by:

$$
\begin{aligned}
\mu_i &= \sum_{k \in K^i} \sum_{j \in L^i_{\hat{k}}, j \neq i} 1 - (1 - q_{i,j}c_{i,j})^d\\
\sigma^2_i &= \sum_{k \in K^i} \sum_{j \in L^i_{\hat{k}}, j \neq i} (1 - q_{i,j}c_{i,j})^d(1 - (1 - q_{i,j}c_{i,j})^d)
\end{aligned}
$$

Heterogeneity in disease transmission was modeled on the one hand by treating the transmission probability as a random variable, and the contact probability on the other hand.

If the individual transmission probability is modeled as a random variable, we can adapt the theoretical description as follows. We assume here that $q$ is determined only by the infectious individual and is not dependent on the (susceptible) individual that this first individual has a contact with. As such, we will henceforth refer to $q$ as $q_i$. Let $q_i$ be a continuous random variable with support on $(0, 1]$ and let $f_q(x)$ be its probability density function. Then the mean number of secondary cases individual $i$ makes during their infectious period can be expressed as:

$$\mu_i = \mathbb{E}\left[\mathbb{E}[Y^i|q_i]\right] = \int_0^\infty \Big( \sum_{k \in K^i} \sum_{j \in L^i_k, j \neq i} 1 - (1 - xc_{i,j})^d f_q(x) dx \Big)$$

Assuming that $q$ follows a Truncated Gamma distribution defined on $(0, 1]$ we obtain:

$$\bar{\mu}_i = \mathbb{E}\left[\mathbb{E}[Y^i|q_i]\right] = \int_0^1 \Big( \sum_{k \in K^i} \sum_{j \in L^i_k, j \neq i} 1 - (1 - xc_{i,j})^d \frac{g(x)}{G(1) - G(0)} dx \Big)$$

Where $g(x), G(x)$ are respectively the density and the cumulative distribution function of the considered Gamma distribution.
The variance:

$$\begin{aligned}\sigma_i^2 &= Var(Y^i) = \mathbb{E}[Var(Y^i|q)] + Var(\mathbb{E}[Y^i|q]) = \\ &= \int_0^1 \Big( \sum_{k \in K^i} \sum_{j \in L^i_k, j \neq i} (1 - xc_{i,j})^d (1 - (1 - xc_{i,j})^d) \frac{g(x)}{G(1) - G(0)} dx \Big) + \\ &+ \int_0^1 \Big( \sum_{k \in K^i} \sum_{j \in L^i_k, j \neq i} 1 - (1 - xc_{i,j})^d - \bar{\mu}_i \Big)^2 \frac{g(x)}{G(1) - G(0)} dx\end{aligned}$$

Conversely, when the individual contact factor is treated as a random variable, this can be expressed as:

$$\mu_i = \mathbb{E}\left[\mathbb{E}[Y^i|cf_{i,j}]\right] = \sum_{k \in K^i} \sum_{j \in L^i_k, j \neq i} \int_0^\infty \Big( 1 - (1 - q_{i,j}(y_{i,j}))^d f_{y_{i,j}}(y_{i,j}) dy_{i,j} \Big)$$

where $y_{i,j} = xc_{i,j}$ is the random variable that describes the contact rate between $i$ and $j$.

##### Simulation

We compared the estimations calculated using the theoretical description to results obtained by simulation. In order to keep the theoretical description tractable, we introduced some simplification to the simulations that we ran to compare to the theoretical description.

First, we used a constant infectious period of 7 days. Secondly, symptomatic cases did not remain in their household, but kept having the same contact rates as before infection. Finally, we treated each simulation day as a weekday – so no weekends or holidays were modeled in these simulations.

We performed these simulations for different individuals: an adult (aged 46 years), a child (aged 5 years) and an elderly person (aged 75 years). We tested the following mean transmission probabilities: 0.025, 0.050, 0.075 and 0.10. As for our other experiments, we tested several values for $\alpha_i$ (10, 1, 0.6, 0.4, 0.2) as well as for $\alpha_c$ (10, 1, 0.6, 0.4, 0.2). We ran each simulation for 40 days, after which the index case should have recovered.

For each scenario, we ran 200 simulations.

##### Results

We compared theoretical estimates with simulation results for the mean and variance of the number of secondary cases per index case for an adult (see Supplementary

Figures S3–S6), a child (see Supplementary Figures S7–S10) and an elderly person (see Supplementary Figures S11–S14).

In all cases, we found that means estimated using the theoretical description closely matched the means of the simulation results. For the variance, the theoretically estimated quantities did not exactly match the simulated ones, but in both results a similar trend could be distinguished.

### S3 Sensitivity analysis

To conduct the sensitivity analysis, we varied the value of two parameters: the number of infected seeds at the beginning of the simulation and the mean transmission probability. In our main analysis, we introduced one infected individual in the population at the beginning of each simulation. For the sensitivity analysis we introduced 10, 50 and 100 infected seeds at the beginning of the simulation. For the mean transmission probability we used 0.08 in our main analysis, and additionally tested the values 0.06 and 0.10 for our sensitivity analysis.

We then checked the behavior of the following outcomes for the scenarios without interventions: the extinction probability (Supplementary Figure S15), the mean attack rate after 200 days (Supplementary Figure S16), the mean size of the peak (Supplementary Figure S17), the mean day on which the peak is reached (Supplementary Figure S18), the mean estimated herd immunity threshold (Supplementary Figure S19), the mean day on which the herd immunity threshold is reached (Supplementary Figure S20), and the mean day on which the last transmission event is observed (Supplementary Figure S21).

We found that, for most outcomes, varying either the number of infected seeds at the beginning of the simulation or the mean transmission probability, led to the same trends being observed when $\alpha_i$ or $\alpha_c$ decreased.

The only exception we observed, was that, with a mean transmission probability of 0.1, the mean size of the peak decreased slightly (instead of increasing) when $\alpha_c$ was decreased from 10 to 0.2 (see Supplementary Figure S17d).

### S4 Supplementary figures

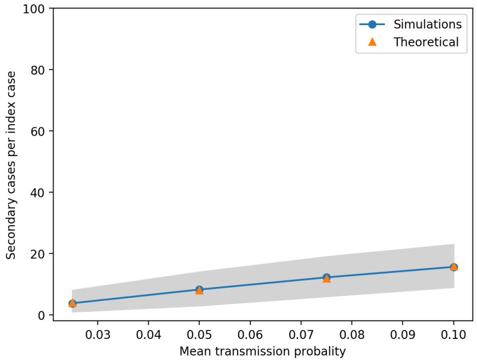


**(a)** Baseline

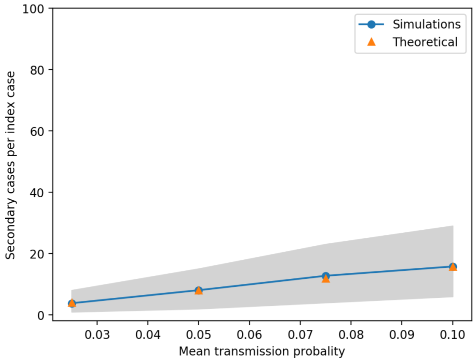


**(b)** $\alpha_i = 10$

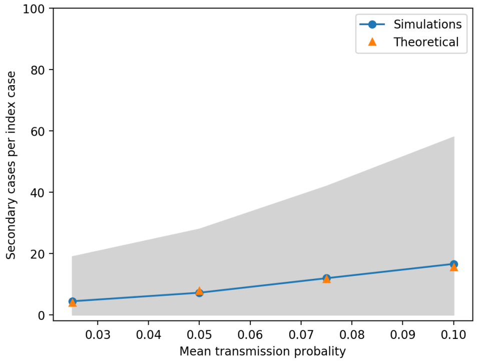


**(c)** $\alpha_i = 1$

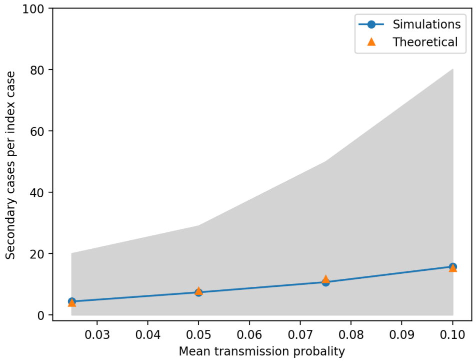


**(d)** $\alpha_i = 0.6$

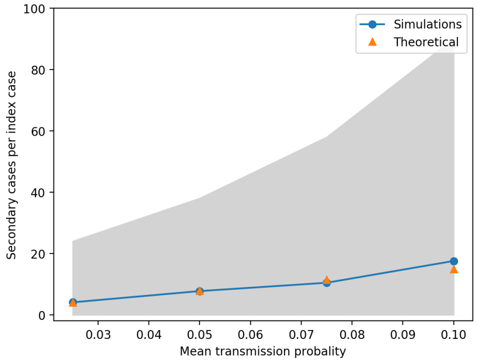


**(e)** $\alpha_i = 0.4$

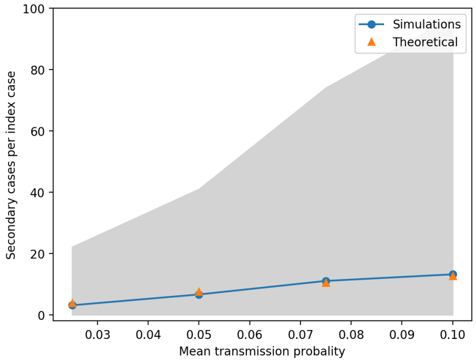


**(f)** $\alpha_i = 0.2$

**Fig S3.** Comparison of the theoretical estimate for the mean individual reproduction number of an adult and the mean number of secondary cases per index case from simulations for the same individual. Different values of $\alpha_i$ for the Truncated Gamma distribution considered for the individual transmission probability were tested.

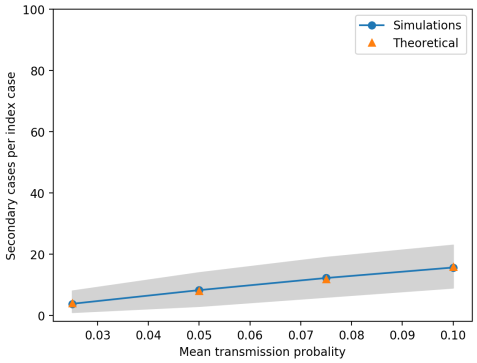


**(a)** Baseline

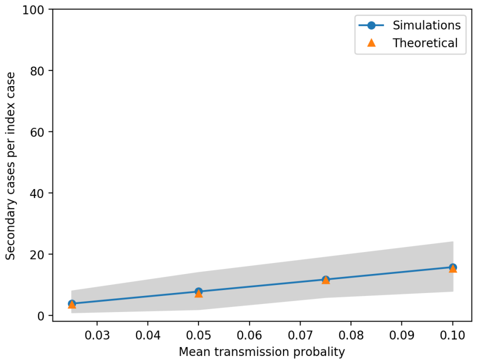


**(b)** $\alpha_c = 10$

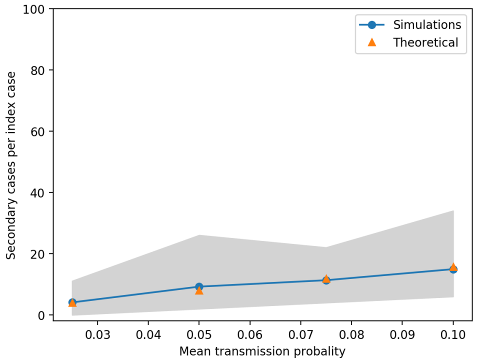


**(c)** $\alpha_c = 1$

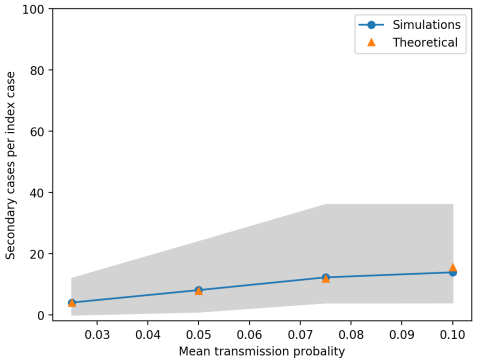


**(d)** $\alpha_c = 0.6$

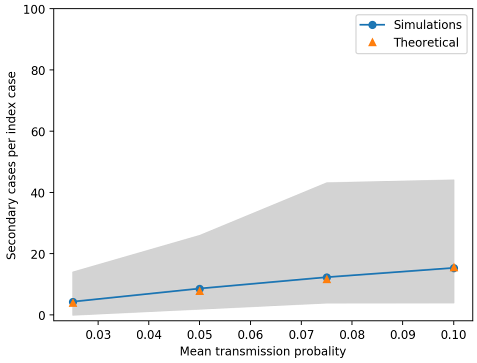


**(e)** $\alpha_c = 0.4$

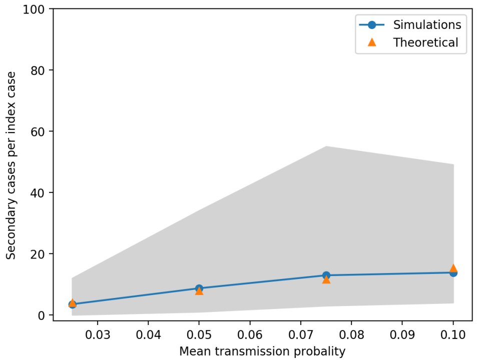


**(f)** $\alpha_c = 0.2$

**Fig S4.** Comparison of the theoretical estimate for the mean individual reproduction number of an adult and the mean number of secondary cases per index case from simulations for the same individual. Different values of $\alpha_c$ for the Gamma distribution considered for the individual contact factor were tested.

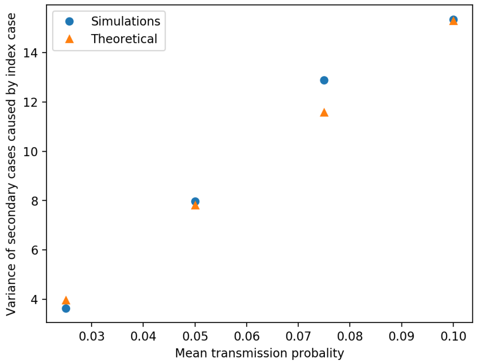


**(a)** Baseline

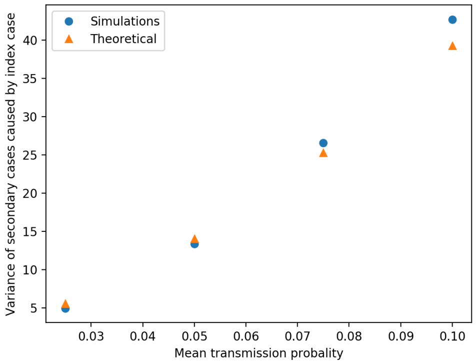


**(b)** $\alpha_i = 10$

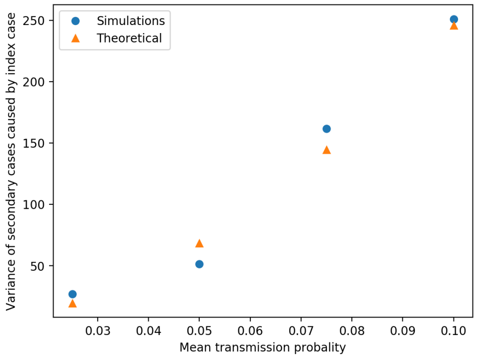


**(c)** $\alpha_i = 1$

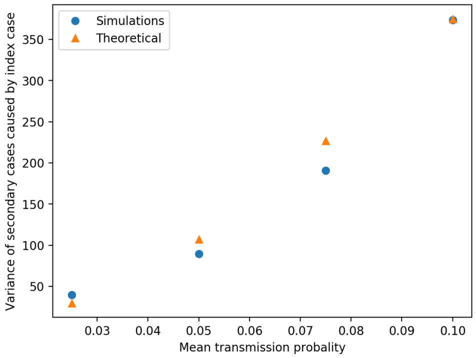


**(d)** $\alpha_i = 0.6$

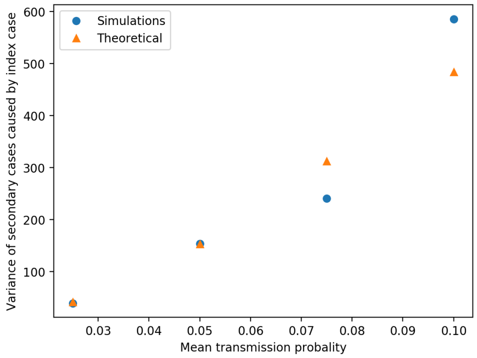


**(e)** $\alpha_i = 0.4$

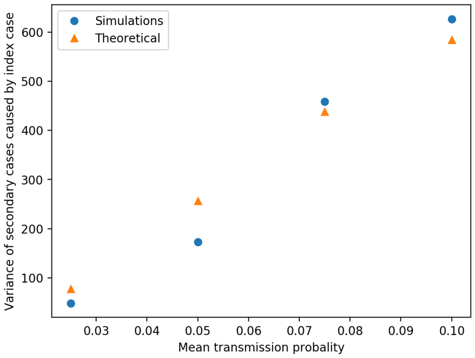


**(f)** $\alpha_i = 0.2$

**Fig S5.** Comparison of the theoretical estimate for the variance of the individual reproduction number of an adult and the variance of the number of secondary cases per index case from simulations for the same individual. Different values of $\alpha_i$ for the Truncated Gamma distribution considered for the individual transmission probability were tested.

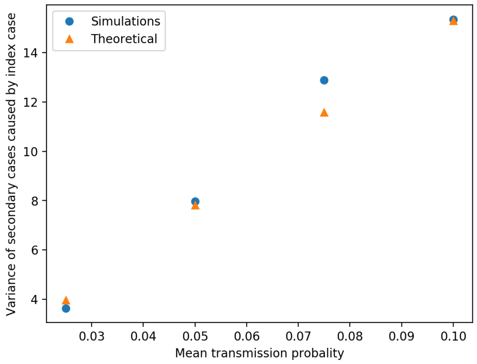


**(a)** Baseline

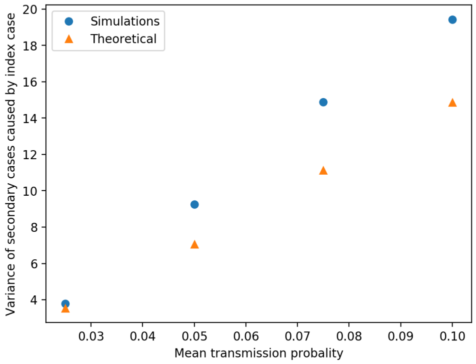


**(b)** $\alpha_i = 10$

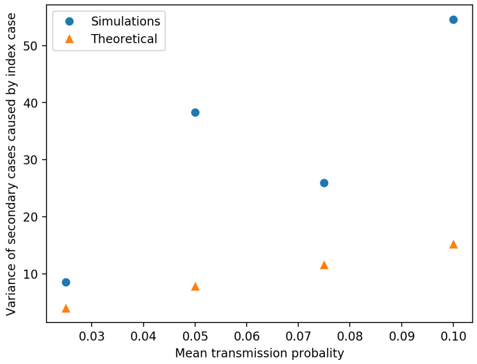


**(c)** $\alpha_i = 1$

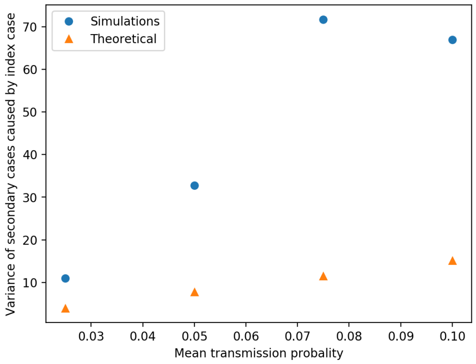


**(d)** $\alpha_i = 0.6$

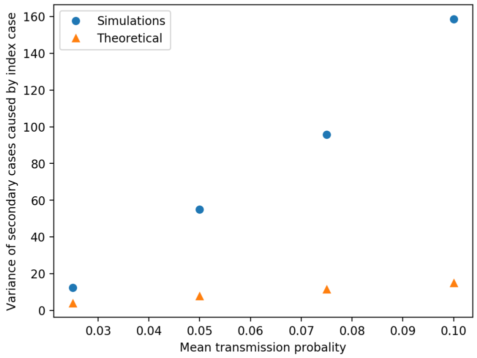


**(e)** $\alpha_i = 0.4$

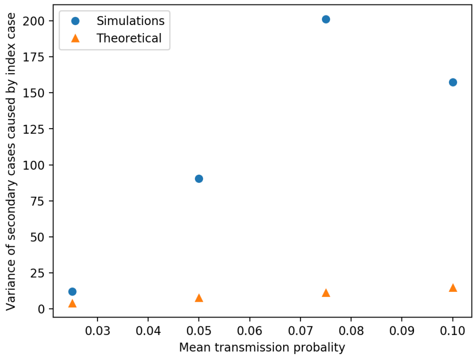


**(f)** $\alpha_i = 0.2$

**Fig S6.** Comparison of the theoretical estimate for the variance of the individual reproduction number of an adult and the variance of the number of secondary cases per index case from simulations for the same individual. Different values of $\alpha_c$ for the Gamma distribution considered for the individual contact factor were tested.

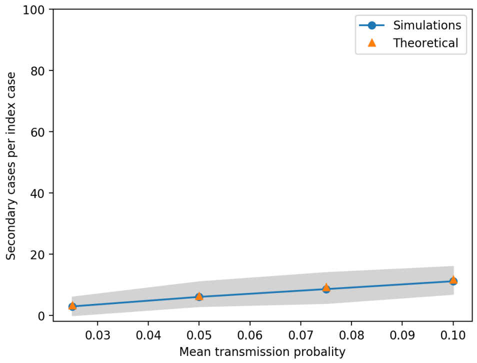


**(a)** Baseline

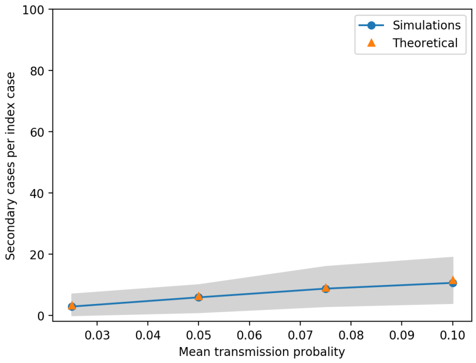


**(b)** $\alpha_i = 10$

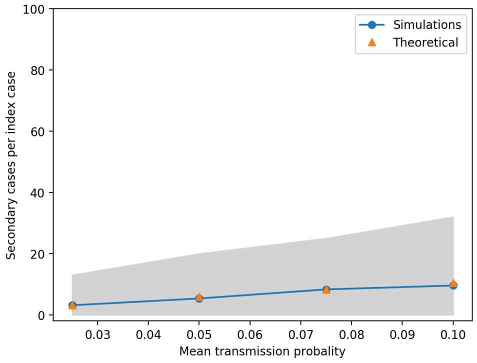


**(c)** $\alpha_i = 1$

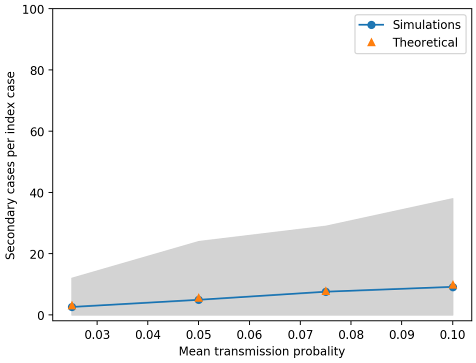


**(d)** $\alpha_i = 0.6$

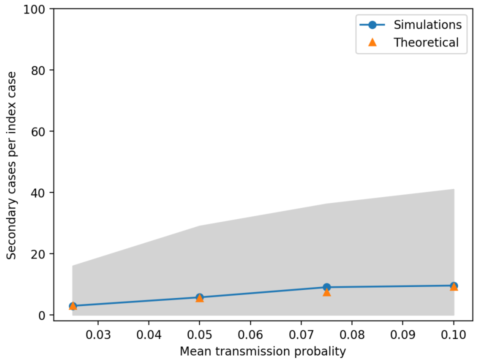


**(e)** $\alpha_i = 0.4$

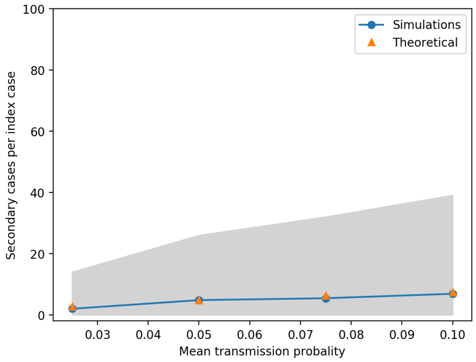


**(f)** $\alpha_i = 0.2$

**Fig S7.** Comparison of the theoretical estimate for the mean individual reproduction number of a child and the mean number of secondary cases per index case from simulations for the same individual. Different values of $\alpha_i$ for the Truncated Gamma distribution considered for the individual transmission probability were tested.

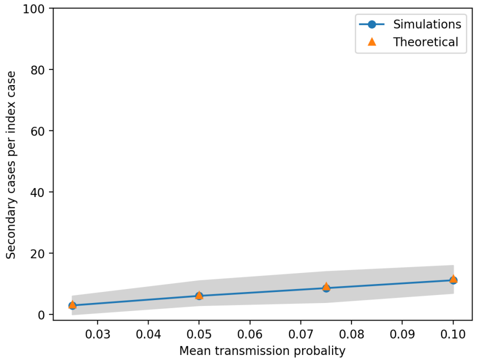


**(a)** Baseline

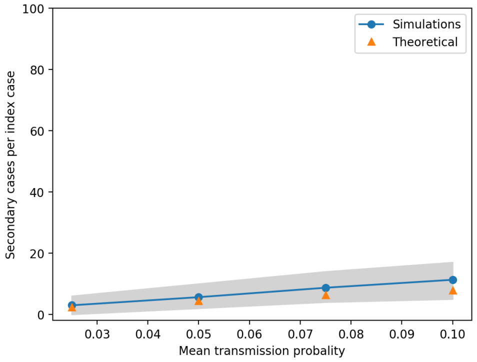


**(b)** $\alpha_c = 10$

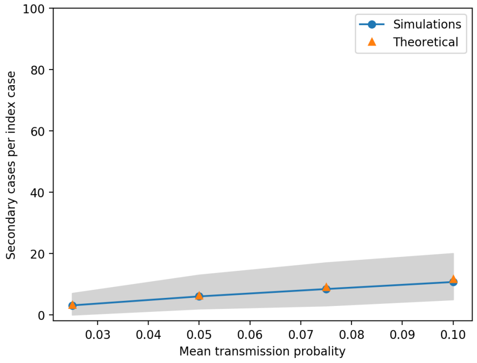


**(c)** $\alpha_c = 1$

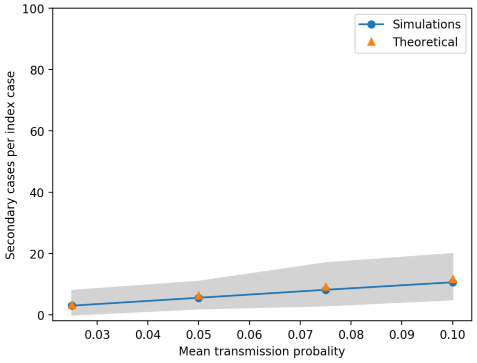


**(d)** $\alpha_c = 0.6$

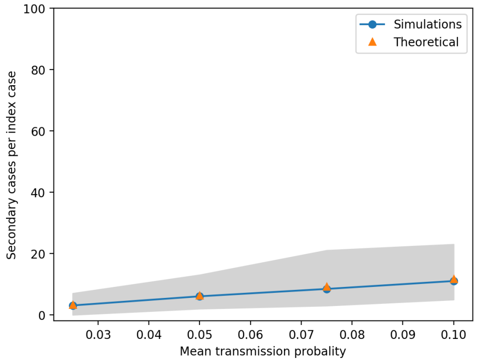


**(e)** $\alpha_c = 0.4$

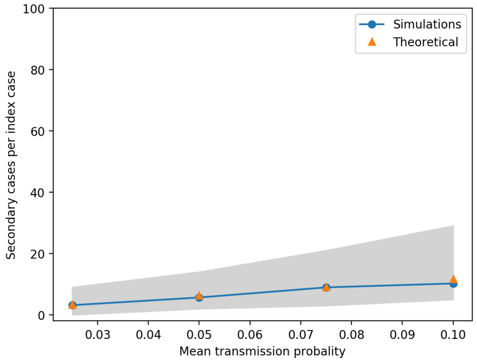


**(f)** $\alpha_c = 0.2$

**Fig S8.** Comparison of the theoretical estimate for the mean individual reproduction number of a child and the mean number of secondary cases per index case from simulations for the same individual. Different values of $\alpha_c$ for the Gamma distribution considered for the individual contact factor were tested.

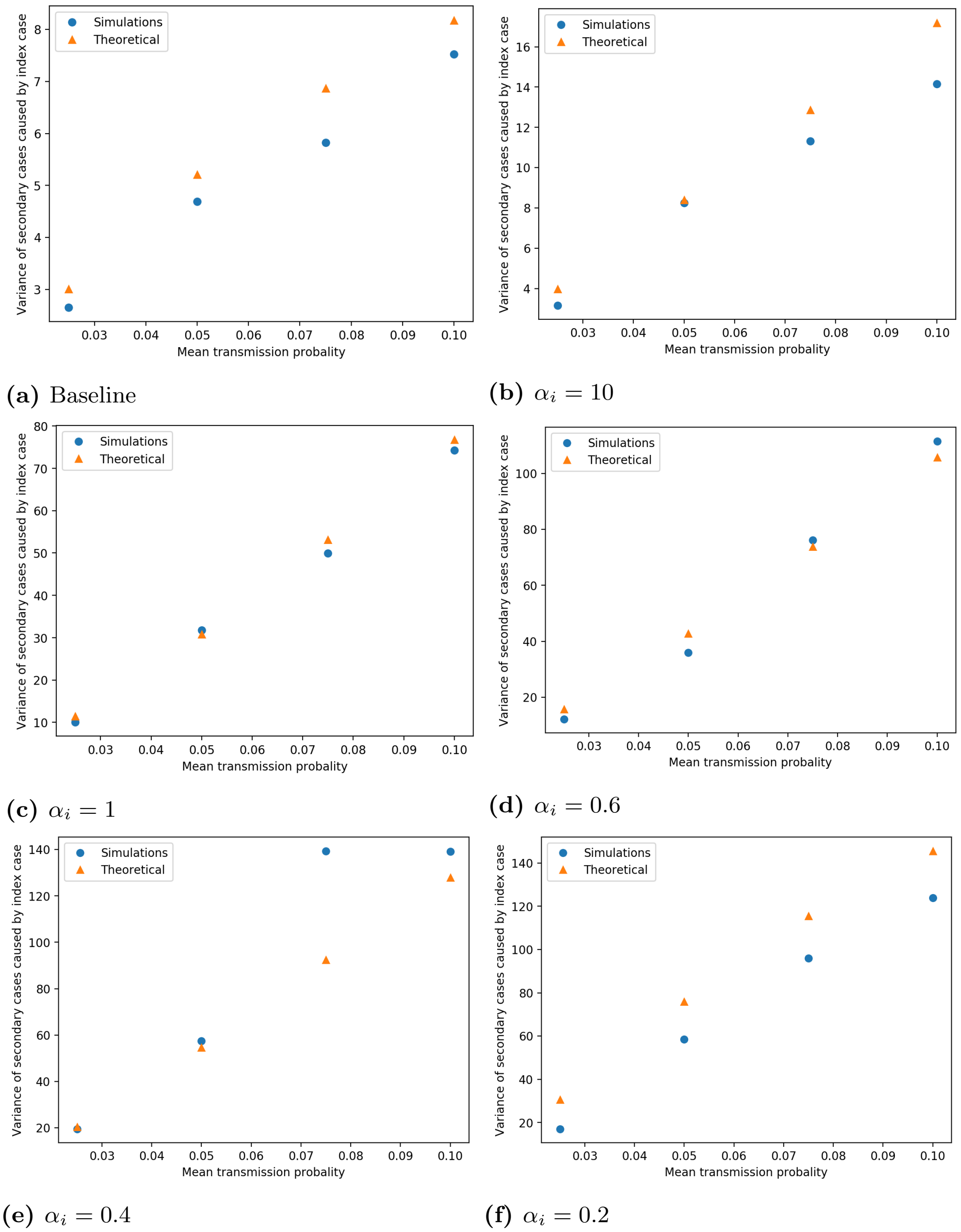


(a) Baseline

(b) $\alpha_i = 10$

(c) $\alpha_i = 1$

(d) $\alpha_i = 0.6$

(e) $\alpha_i = 0.4$

(f) $\alpha_i = 0.2$

**Fig S9.** Comparison of the theoretical estimate for the variance of the individual reproduction number of a child and the variance of the number of secondary cases per index case from simulations for the same individual. Different values of $\alpha_i$ for the Truncated Gamma distribution considered for the individual transmission probability were tested.

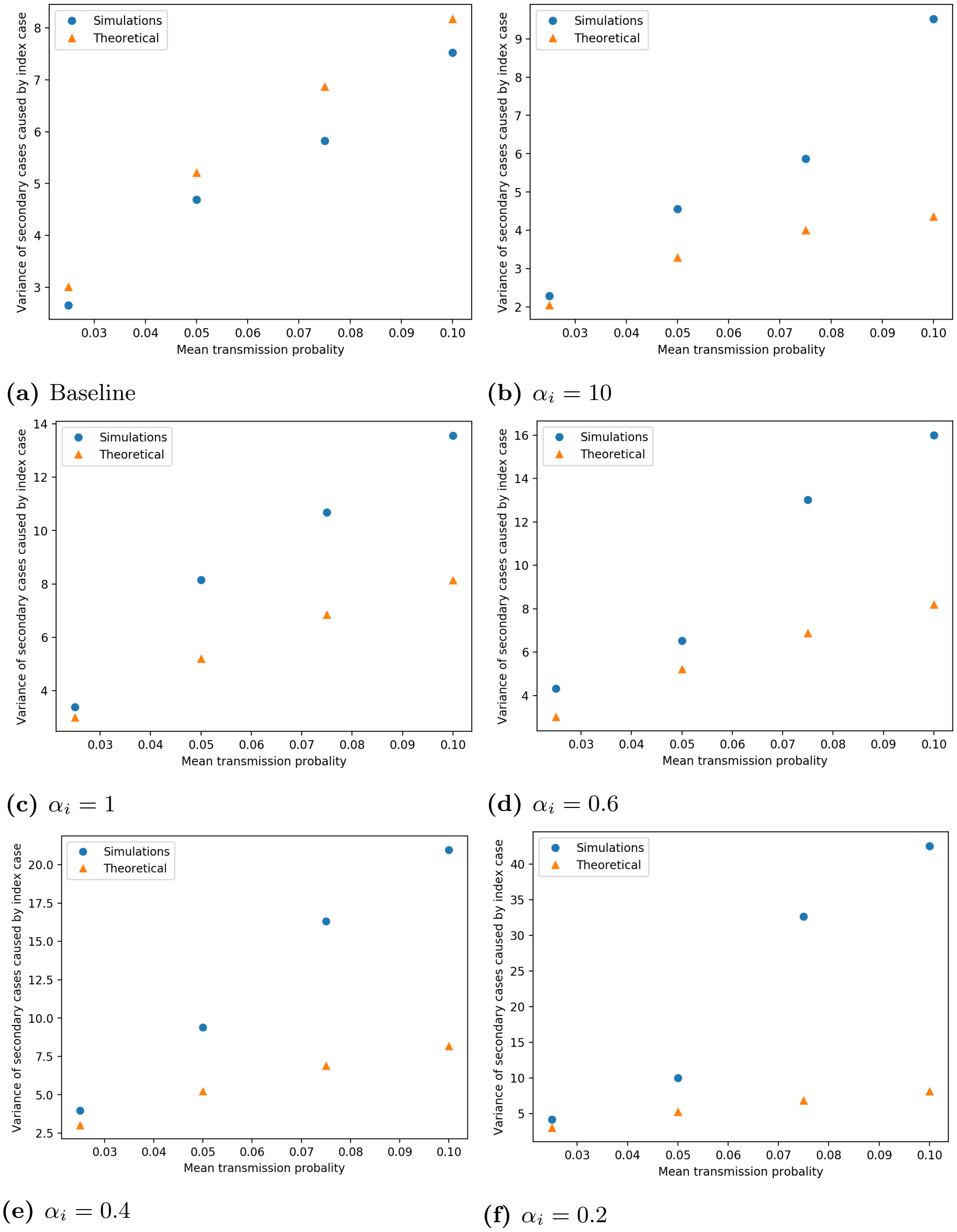


**(a)** Baseline

**(b)** $\alpha_i = 10$

**(c)** $\alpha_i = 1$

**(d)** $\alpha_i = 0.6$

**(e)** $\alpha_i = 0.4$

**(f)** $\alpha_i = 0.2$

**Fig S10.** Comparison of the theoretical estimate for the variance of the individual reproduction number of a child and the variance of the number of secondary cases per index case from simulations for the same individual. Different values of $\alpha_c$ for the Gamma distribution considered for the individual contact factor were tested.

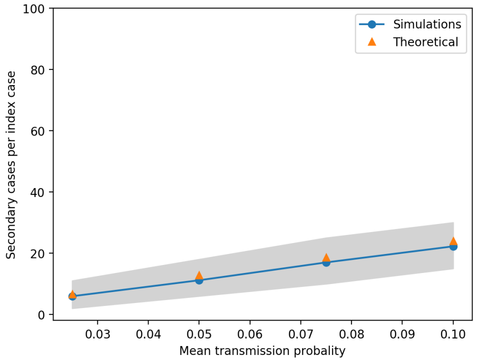


**(a)** Baseline

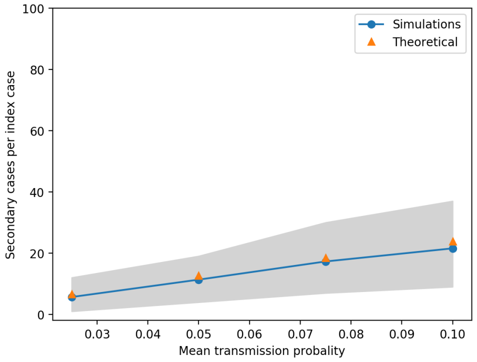


**(b)** $\alpha_i = 10$

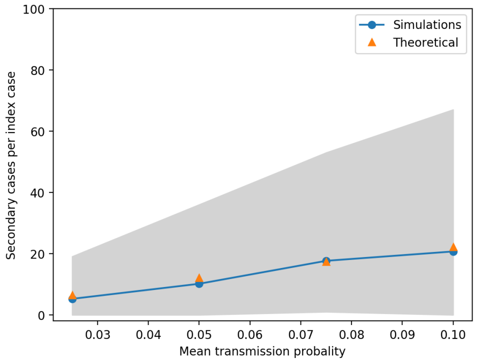


**(c)** $\alpha_i = 1$

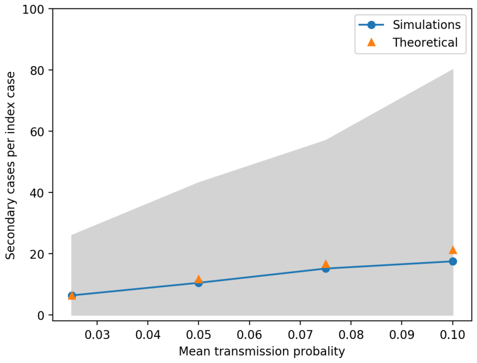


**(d)** $\alpha_i = 0.6$

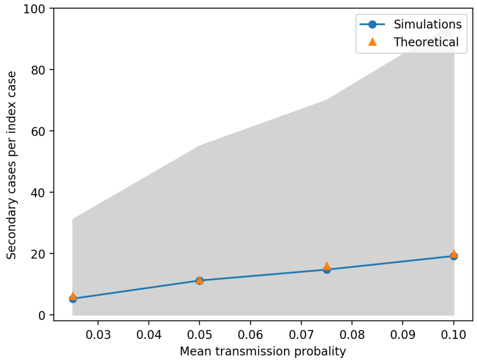


**(e)** $\alpha_i = 0.4$

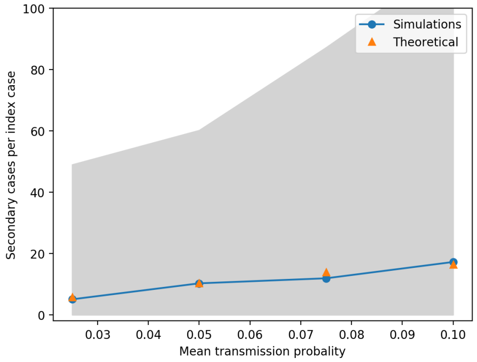


**(f)** $\alpha_i = 0.2$

**Fig S11.** Comparison of the theoretical estimate for the mean individual reproduction number of an elderly person and the mean number of secondary cases per index case from simulations for the same individual. Different values of $\alpha_i$ for the Truncated Gamma distribution considered for the individual transmission probability were tested.

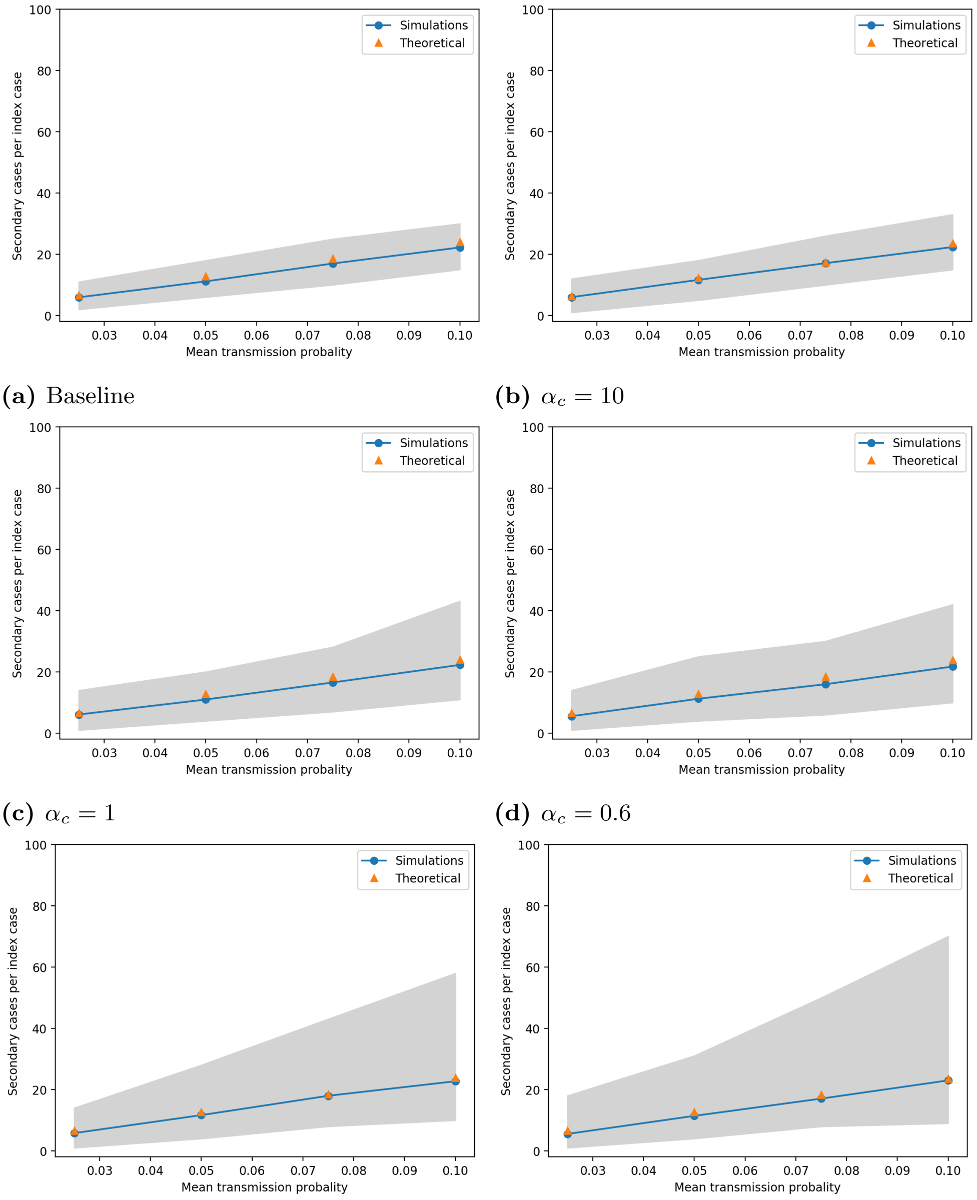


**(a)** Baseline

**(b)** $\alpha_c = 10$

**(c)** $\alpha_c = 1$

**(d)** $\alpha_c = 0.6$

**(e)** $\alpha_c = 0.4$

**(f)** $\alpha_c = 0.2$

**Fig S12.** Comparison of the theoretical estimate for the mean individual reproduction number of an elderly person and the mean number of secondary cases per index case from simulations for the same individual. Different values of $\alpha_c$ for the Gamma distribution considered for the individual contact factor were tested.

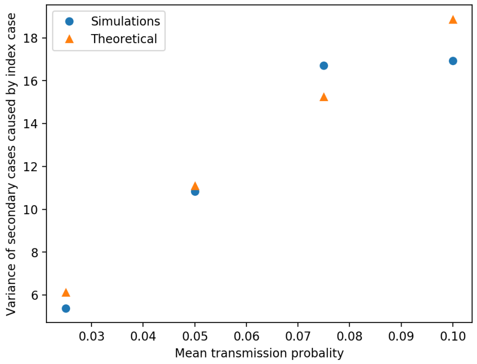


**(a)** Baseline

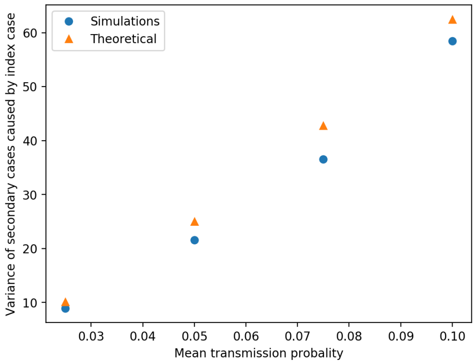


**(b)** $\alpha_i = 10$

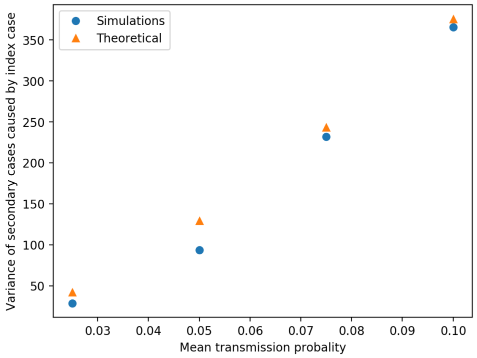


**(c)** $\alpha_i = 1$

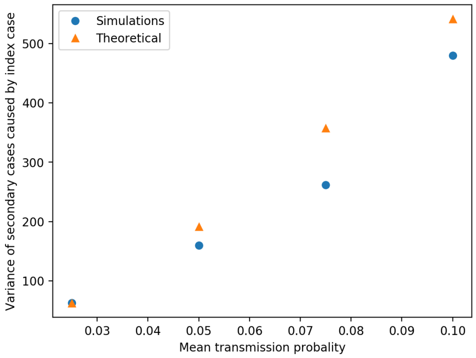


**(d)** $\alpha_i = 0.6$

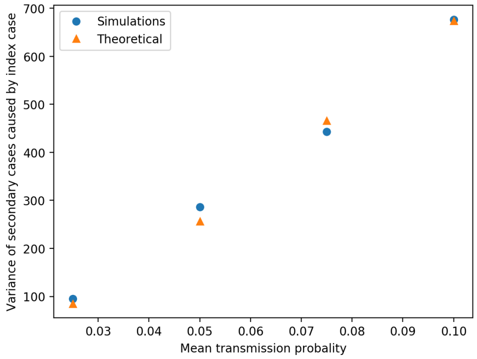


**(e)** $\alpha_i = 0.4$

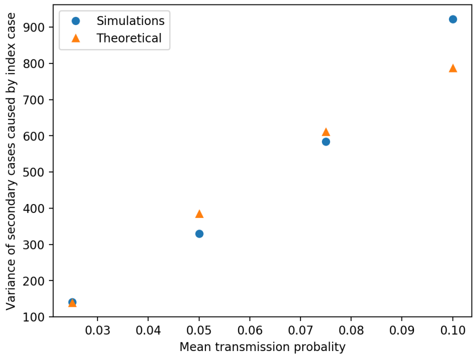


**(f)** $\alpha_i = 0.2$

**Fig S13.** Comparison of the theoretical estimate for the variance of the individual reproduction number of an elderly person and the variance of the number of secondary cases per index case from simulations for the same individual. Different values of $\alpha_i$ for the Truncated Gamma distribution considered for the individual transmission probability were tested.

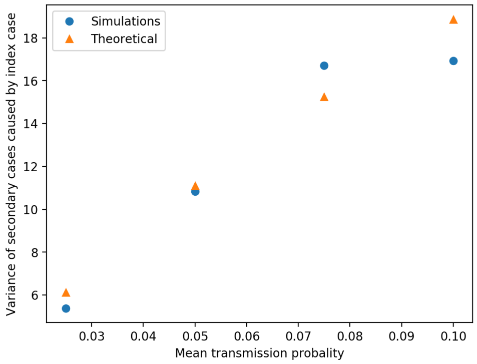


**(a)** Baseline

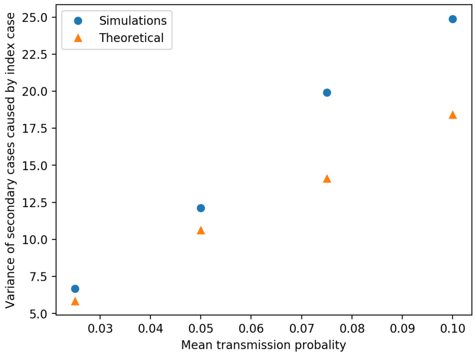


**(b)** $\alpha_i = 10$

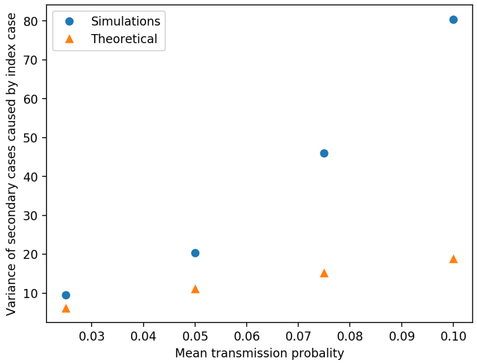


**(c)** $\alpha_i = 1$

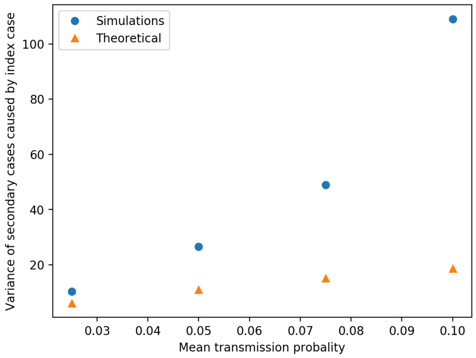


**(d)** $\alpha_i = 0.6$

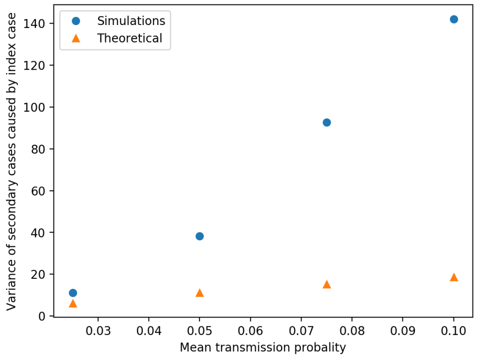


**(e)** $\alpha_i = 0.4$

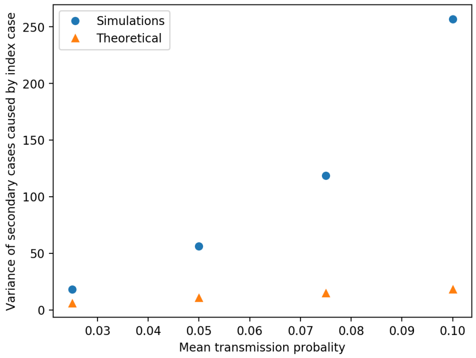


**(f)** $\alpha_i = 0.2$

**Fig S14.** Comparison of the theoretical estimate for the variance of the individual reproduction number of an elderly person and the variance of the number of secondary cases per index case from simulations for the same individual. Different values of $\alpha_c$ for the Gamma distribution considered for the individual contact factor were tested.

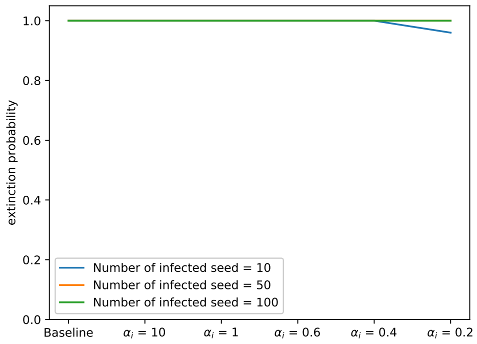


**(a)** Impact of introducing different numbers of infected seeds at the beginning of the simulation when varying $\alpha_i$ for the Truncated Gamma distribution considered for the individual transmission probability.

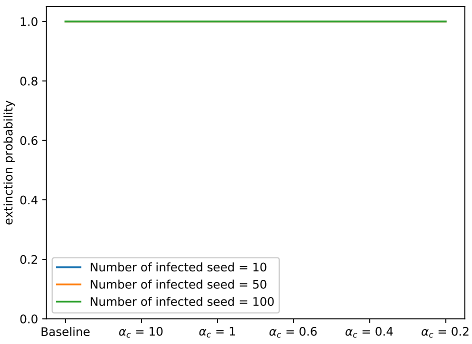


**(b)** Impact of introducing different numbers of infected seeds at the beginning of the simulation when varying $\alpha_c$ for the Gamma distribution considered for the individual contact factor.

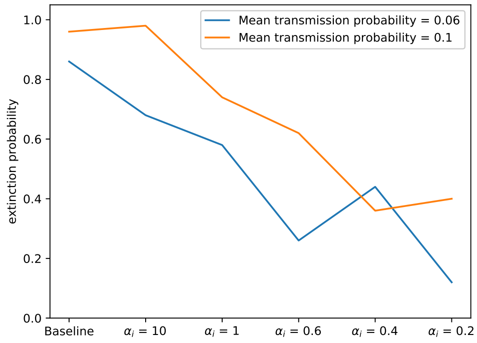


**(c)** Impact of different values for the mean transmission probability when varying $\alpha_i$ for the Truncated Gamma distribution considered for the individual transmission probability.

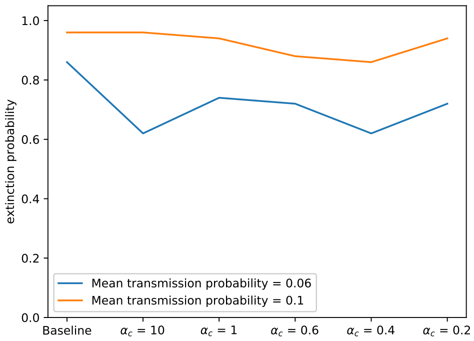


**(d)** Impact of different values for the mean transmission probability when varying $\alpha_c$ for the Gamma distribution considered for the individual contact factor.

**Fig S15.** Impact of different values for the number of infected seeds at the beginning of the simulation (panel a and b) and for the mean transmission probability (panel c and d) on the extinction probability. The extinction threshold was set at 50 cases.

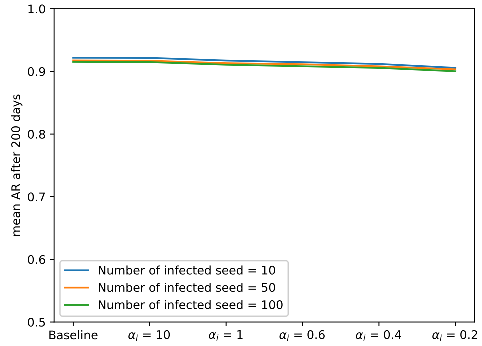


**(a)** Impact of introducing different numbers of infected seeds at the beginning of the simulation when varying $\alpha_i$ for the Truncated Gamma distribution considered for the individual transmission probability.

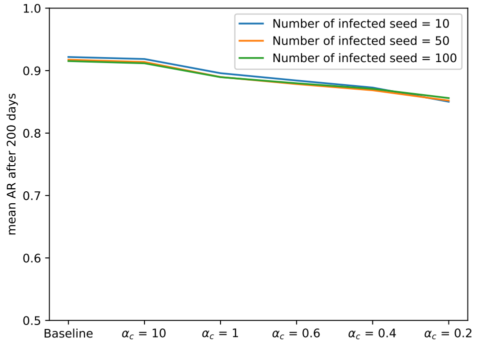


**(b)** Impact of introducing different numbers of infected seeds at the beginning of the simulation when varying $\alpha_c$ for the Gamma distribution considered for the individual contact factor.

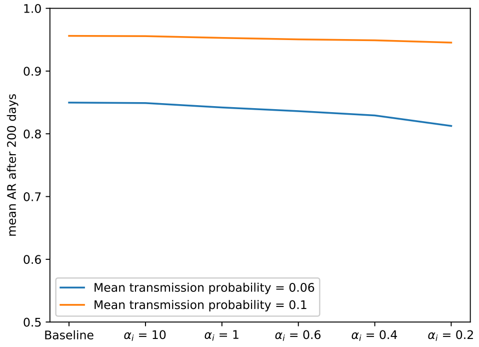


**(c)** Impact of different values for the mean transmission probability when varying $\alpha_i$ for the Truncated Gamma distribution considered for the individual transmission probability.

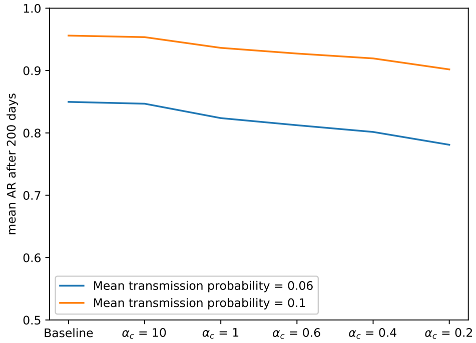


**(d)** Impact of different values for the mean transmission probability when varying $\alpha_c$ for the Gamma distribution considered for the individual contact factor.

**Fig S16.** Impact of different values for the number of infected seeds at the beginning of the simulation (panel a and b) and for the mean transmission probability (panel c and d) on the mean attack rate over 200 days. Simulation runs which led to extinction (i.e. those that produced less than 50 cases) were excluded.

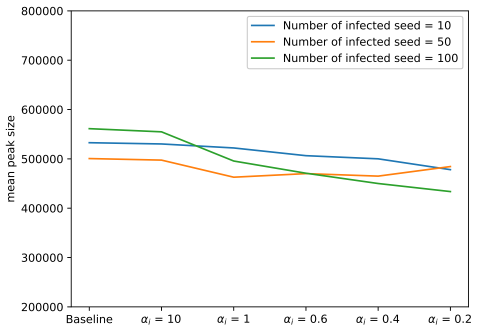


**(a)** Impact of introducing different numbers of infected seeds at the beginning of the simulation when varying $\alpha_i$ for the Truncated Gamma distribution considered for the individual transmission probability.

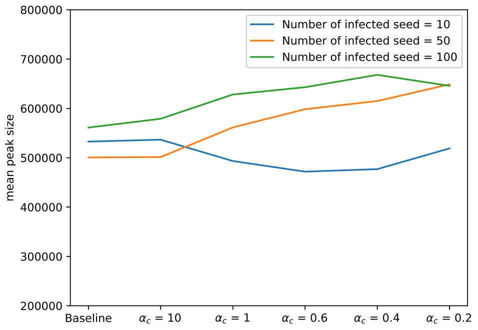


**(b)** Impact of introducing different numbers of infected seeds at the beginning of the simulation when varying $\alpha_c$ for the Gamma distribution considered for the individual contact factor.

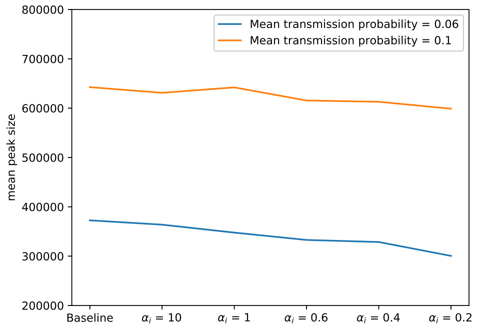


**(c)** Impact of different values for the mean transmission probability when varying $\alpha_i$ for the Truncated Gamma distribution considered for the individual transmission probability.

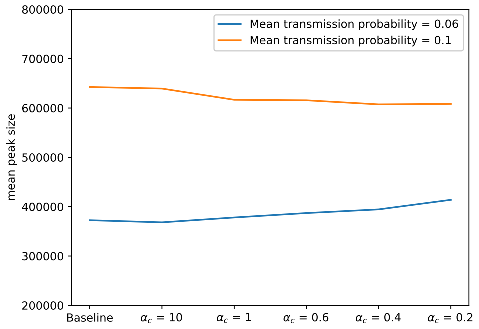


**(d)** Impact of different values for the mean transmission probability when varying $\alpha_c$ for the Gamma distribution considered for the individual contact factor.

**Fig S17.** Impact of different values for the number of infected seeds at the beginning of the simulation (panel a and b) and for the mean transmission probability (panel c and d) on the mean size of the peak. Simulation runs which led to extinction (i.e. those that produced less than 50 cases) were excluded.

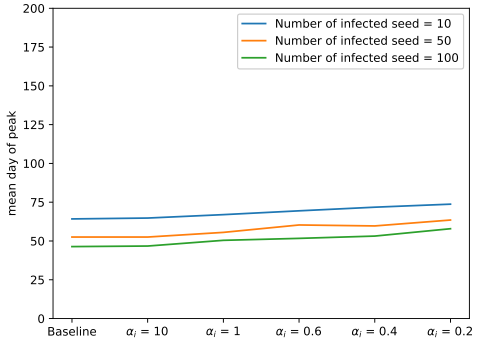


**(a)** Impact of introducing different numbers of infected seeds at the beginning of the simulation when varying $\alpha_i$ for the Truncated Gamma distribution considered for the individual transmission probability.

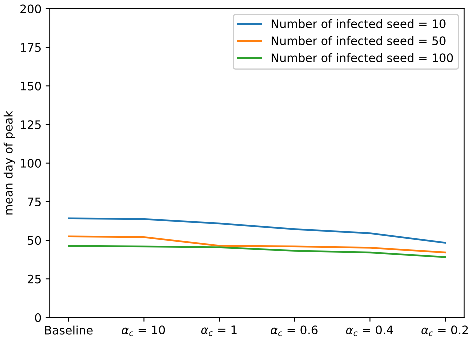


**(b)** Impact of introducing different numbers of infected seeds at the beginning of the simulation when varying $\alpha_c$ for the Gamma distribution considered for the individual contact factor.

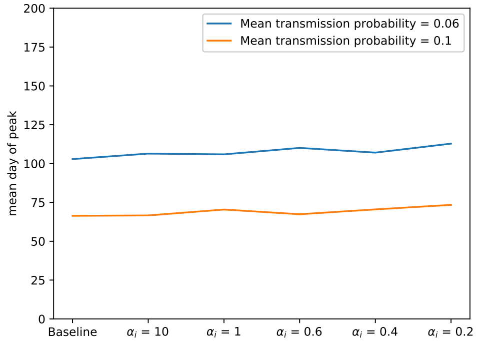


**(c)** Impact of different values for the mean transmission probability when varying $\alpha_i$ for the Truncated Gamma distribution considered for the individual transmission probability.

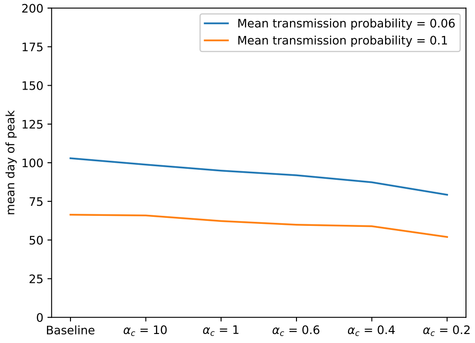


**(d)** Impact of different values for the mean transmission probability when varying $\alpha_c$ for the Gamma distribution considered for the individual contact factor.

**Fig S18.** Impact of different values for the number of infected seeds at the beginning of the simulation (panel a and b) and for the mean transmission probability (panel c and d) on the mean day on which the peak is reached. Simulation runs which led to extinction (i.e. those that produced less than 50 cases) were excluded.

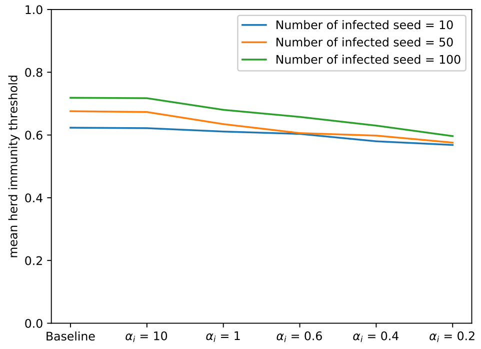


**(a)** Impact of introducing different numbers of infected seeds at the beginning of the simulation when varying $\alpha_i$ for the Truncated Gamma distribution considered for the individual transmission probability.

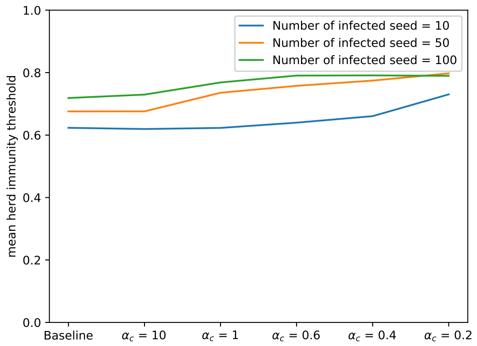


**(b)** Impact of introducing different numbers of infected seeds at the beginning of the simulation when varying $\alpha_c$ for the Gamma distribution considered for the individual contact factor.

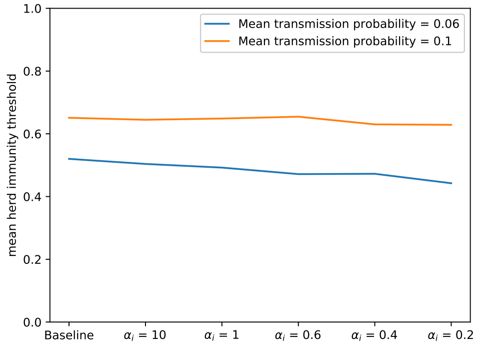


**(c)** Impact of different values for the mean transmission probability when varying $\alpha_i$ for the Truncated Gamma distribution considered for the individual transmission probability.

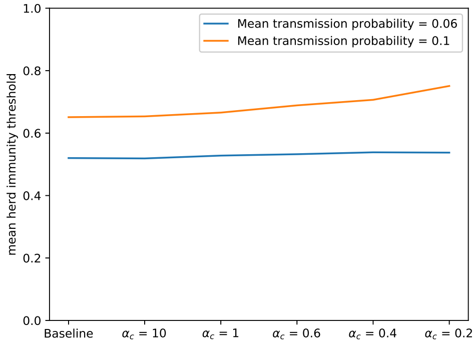


**(d)** Impact of different values for the mean transmission probability when varying $\alpha_c$ for the Gamma distribution considered for the individual contact factor.

**Fig S19.** Impact of different values for the number of infected seeds at the beginning of the simulation (panel a and b) and for the mean transmission probability (panel c and d) on the mean estimated herd immunity threshold. Simulation runs which led to extinction (i.e. those that produced less than 50 cases) were excluded.

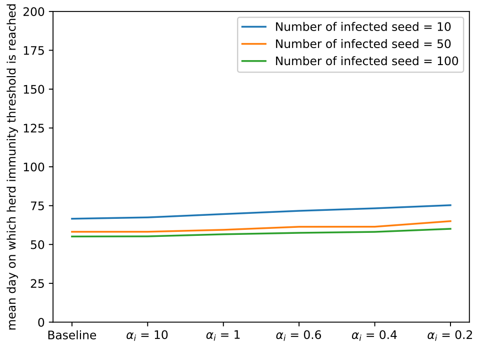


**(a)** Impact of introducing different numbers of infected seeds at the beginning of the simulation when varying $\alpha_i$ for the Truncated Gamma distribution considered for the individual transmission probability.

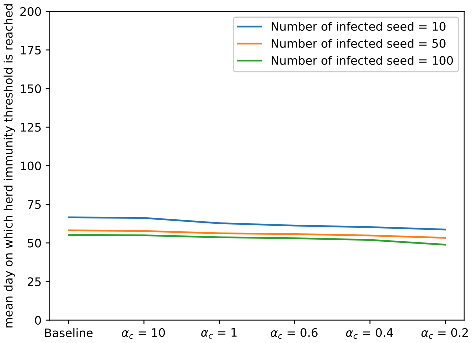


**(b)** Impact of introducing different numbers of infected seeds at the beginning of the simulation when varying $\alpha_c$ for the Gamma distribution considered for the individual contact factor.

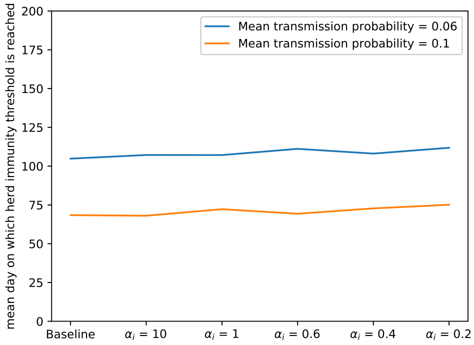


**(c)** Impact of different values for the mean transmission probability when varying $\alpha_i$ for the Truncated Gamma distribution considered for the individual transmission probability.

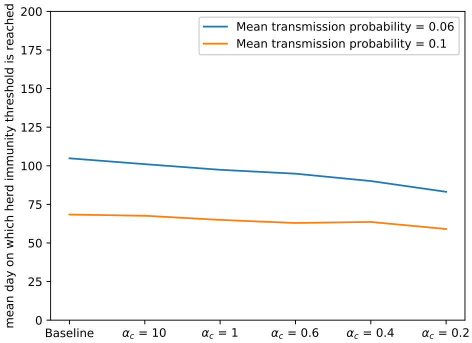


**(d)** Impact of different values for the mean transmission probability when varying $\alpha_c$ for the Gamma distribution considered for the individual contact factor.

**Fig S20.** Impact of different values for the number of infected seeds at the beginning of the simulation (panel a and b) and for the mean transmission probability (panel c and d) on the mean day on which the herd immunity threshold is reached. Simulation runs which led to extinction (i.e. those that produced less than 50 cases) were excluded.

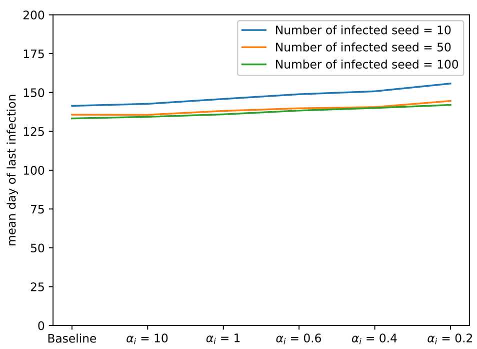


**(a)** Impact of introducing different numbers of infected seeds at the beginning of the simulation when varying $\alpha_i$ for the Truncated Gamma distribution considered for the individual transmission probability.

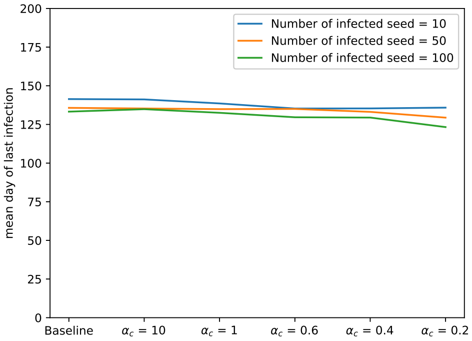


**(b)** Impact of introducing different numbers of infected seeds at the beginning of the simulation when varying $\alpha_c$ for the Gamma distribution considered for the individual contact factor.

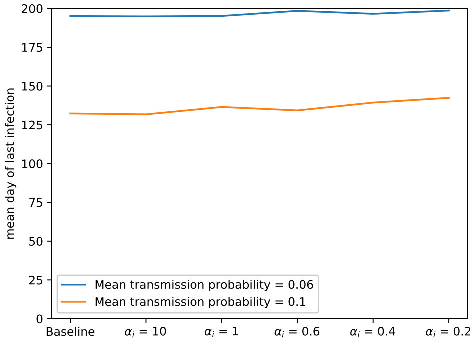


**(c)** Impact of different values for the mean transmission probability when varying $\alpha_i$ for the Truncated Gamma distribution considered for the individual transmission probability.

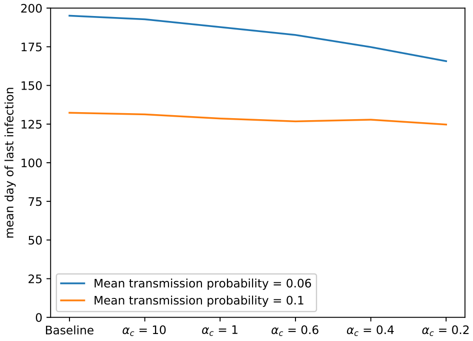


**(d)** Impact of different values for the mean transmission probability when varying $\alpha_c$ for the Gamma distribution considered for the individual contact factor.

**Fig S21.** Impact of different values for the number of infected seeds at the beginning of the simulation (panel a and b) and for the mean transmission probability (panel c and d) on the mean day on which the last transmission event is observed. Simulation runs which led to extinction (i.e. those that produced less than 50 cases) were excluded.

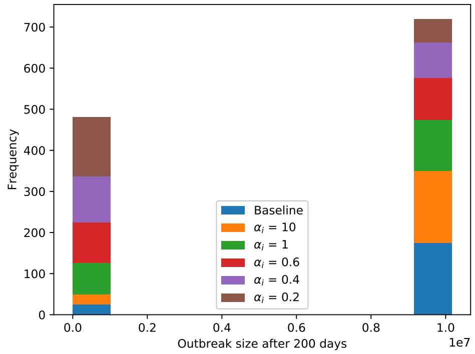


**(a)** Varying $\alpha_i$ for the Truncated Gamma distribution considered the individual transmission probability.

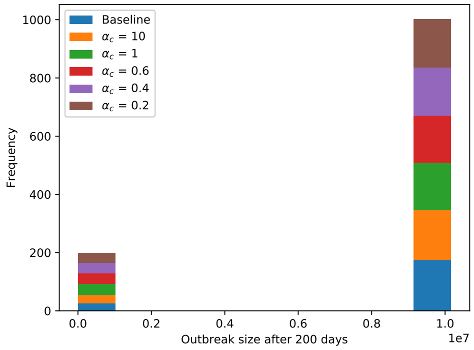


**(b)** Varying $\alpha_c$ for the Gamma distribution considered for the individual contact factor.

**Fig S22.** Histograms of final sizes for the different scenarios regarding infectiousness-related and contact-related heterogeneity, without interventions.

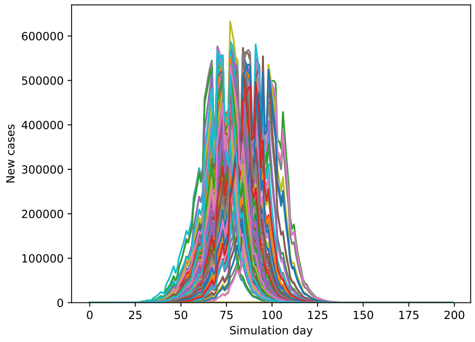


**(a)** Baseline

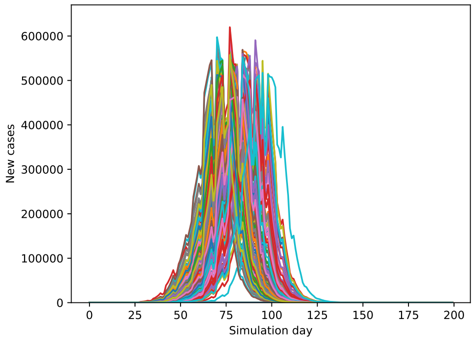


**(b)** $\alpha_i = 10$

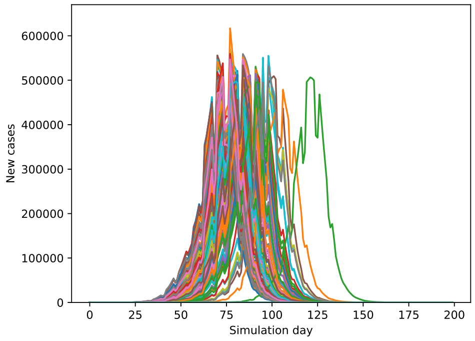


**(c)** $\alpha_i = 1$

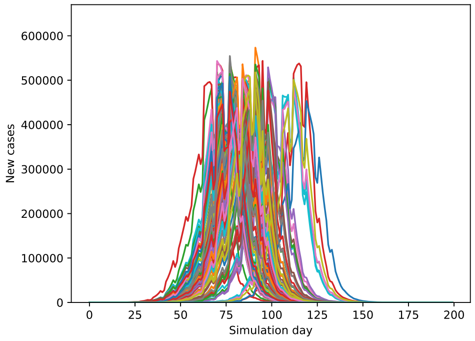


**(d)** $\alpha_i = 0.6$

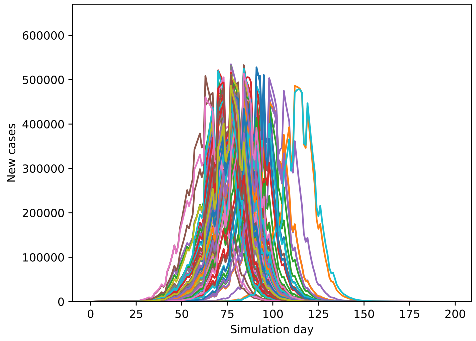


**(e)** $\alpha_i = 0.4$

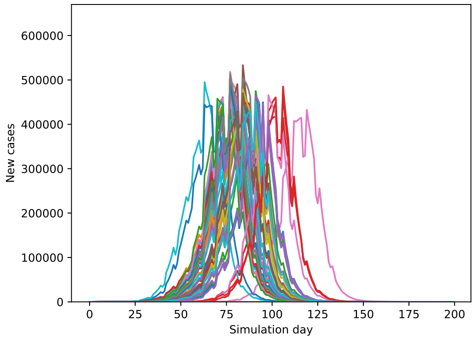


**(f)** $\alpha_i = 0.2$

**Fig S23.** Evolution of the number of new cases per day for different values of $\alpha_i$ for the Truncated Gamma distribution considered for the individual transmission probability, for the scenario without interventions.

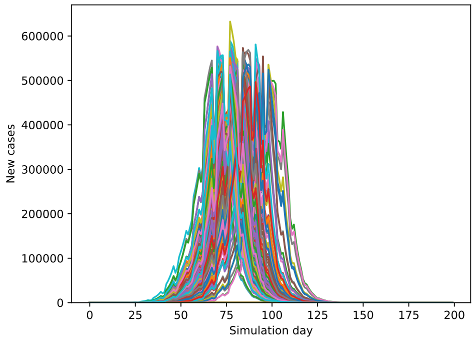


**(a)** Baseline

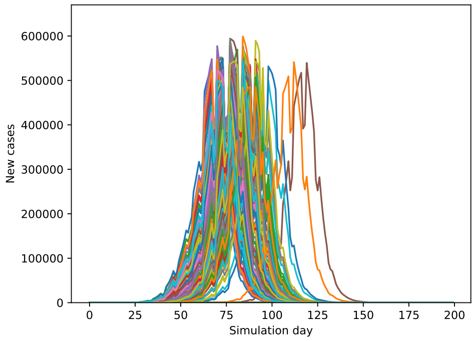


**(b)** $\alpha_c = 10$

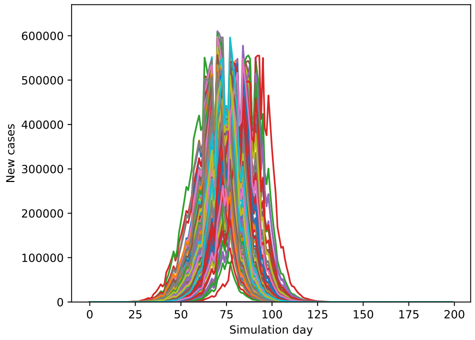


**(c)** $\alpha_c = 1$

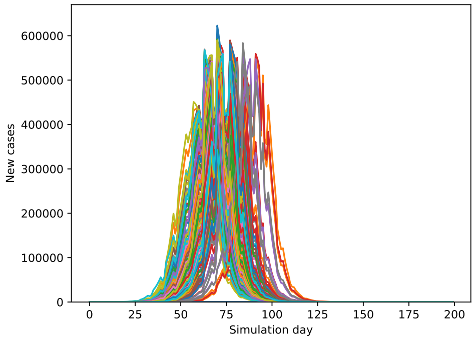


**(d)** $\alpha_c = 0.6$

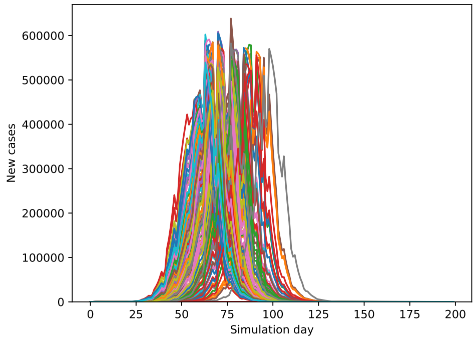


**(e)** $\alpha_c = 0.4$

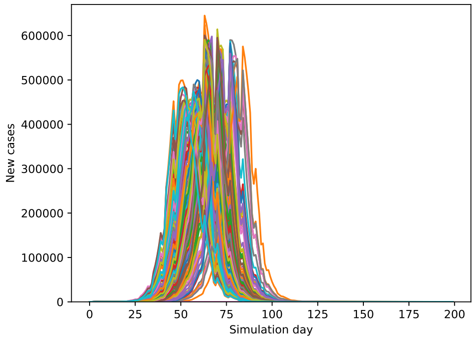


**(f)** $\alpha_c = 0.2$

**Fig S24.** Evolution of the number of new cases per day for different values of $\alpha_c$ for the Gamma distribution considered for the individual contact factor, for the scenario without interventions.

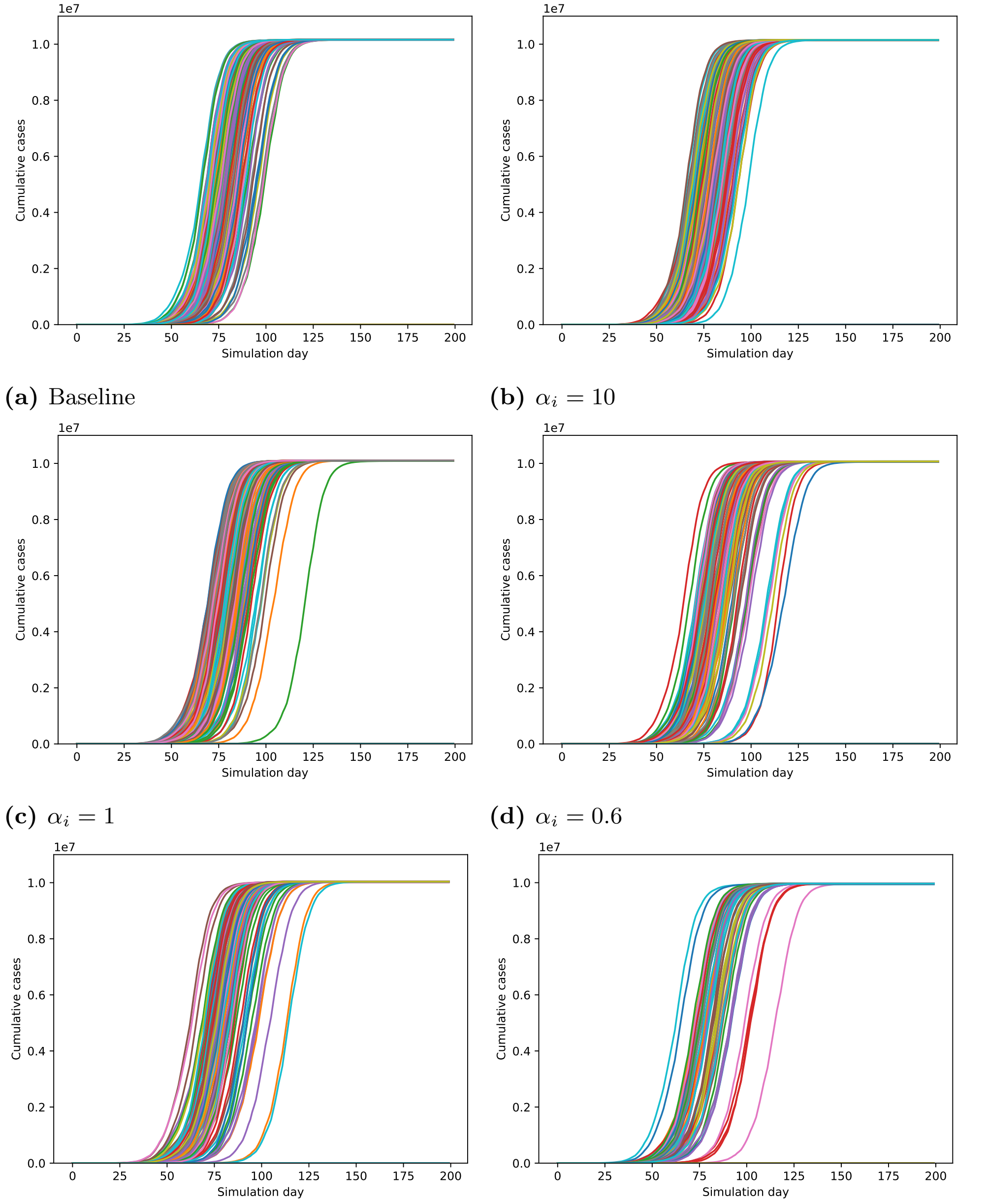


**(a)** Baseline **(b)** $\alpha_i = 10$

**(c)** $\alpha_i = 1$ **(d)** $\alpha_i = 0.6$

**(e)** $\alpha_i = 0.4$ **(f)** $\alpha_i = 0.2$

**Fig S25.** Evolution of the number of cumulative cases per day for different values of $\alpha_i$ for the Truncated Gamma distribution considered for the individual transmission probability, for the scenario without interventions.

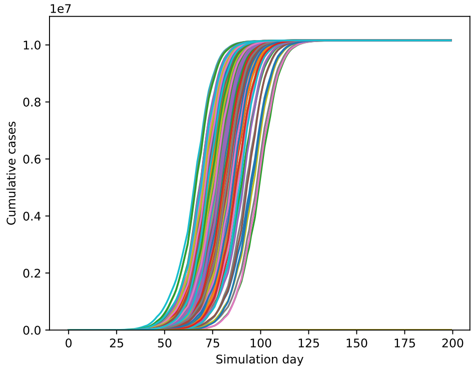


**(a)** Baseline

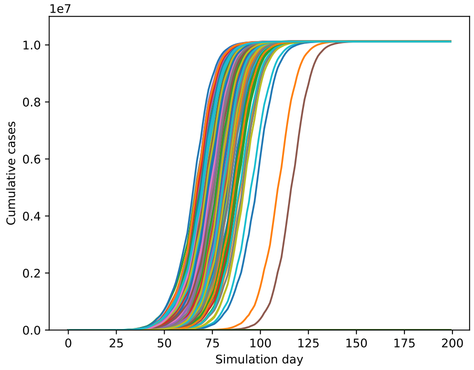


**(b)** $\alpha_c = 10$

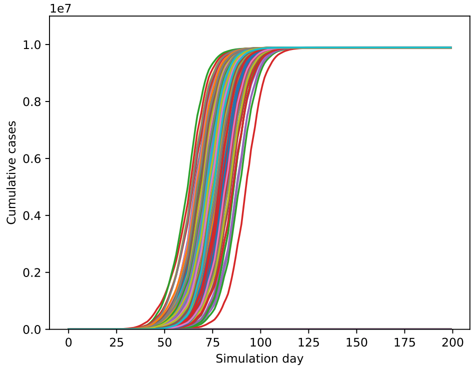


**(c)** $\alpha_c = 1$

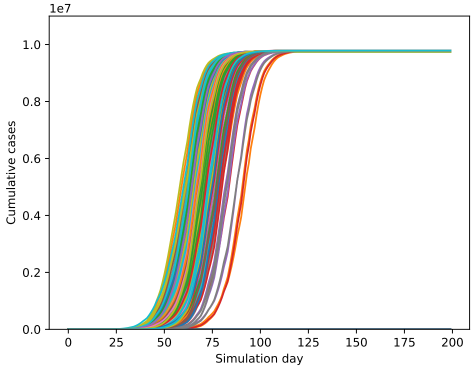


**(d)** $\alpha_c = 0.6$

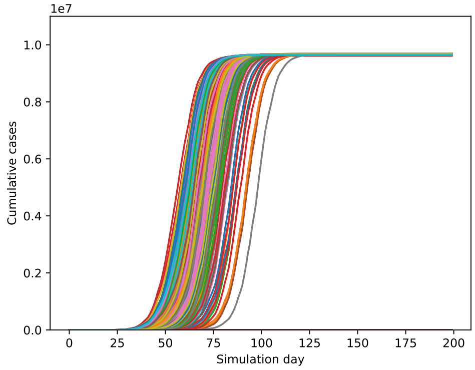


**(e)** $\alpha_c = 0.4$

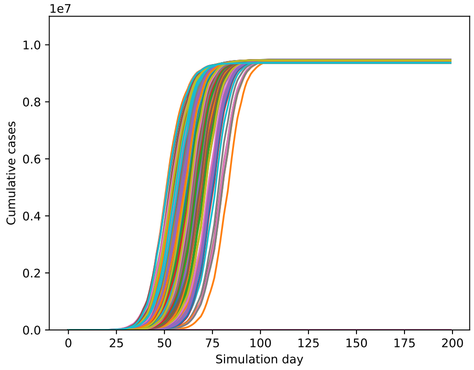


**(f)** $\alpha_c = 0.2$

**Fig S26.** Evolution of the number of cumulative cases per day for different values of $\alpha_c$ for the Gamma distribution considered for the individual contact factor, for the scenario without interventions.

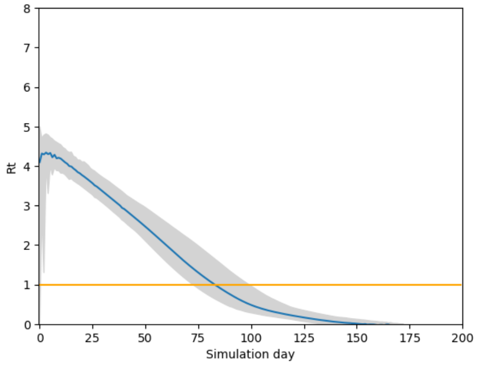


**(a)** Baseline

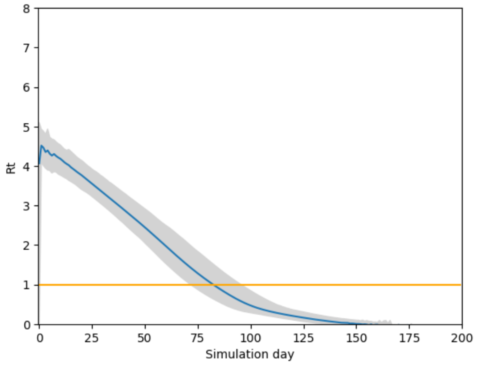


**(b)** $\alpha_i = 10$

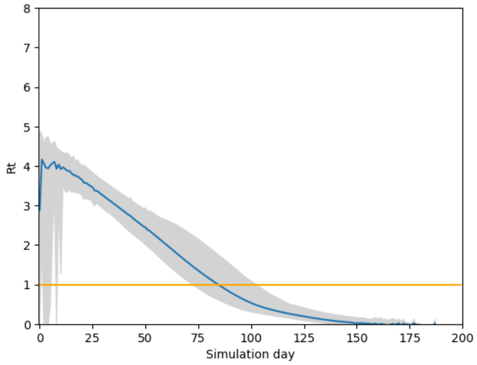


**(c)** $\alpha_i = 1$

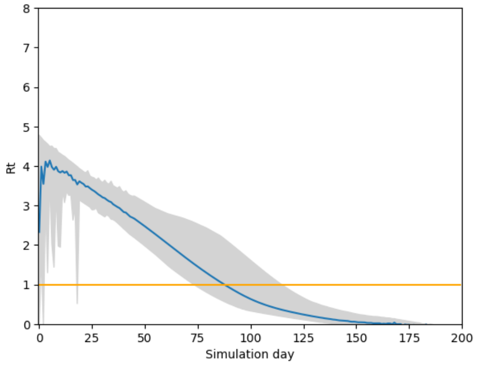


**(d)** $\alpha_i = 0.6$

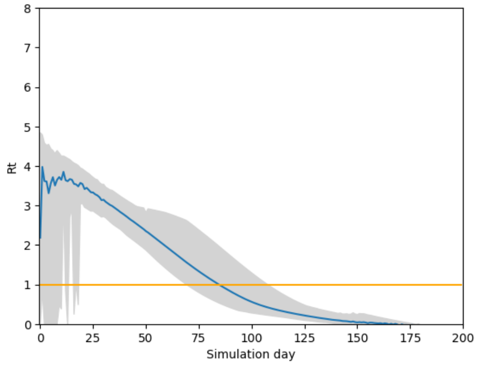


**(e)** $\alpha_i = 0.4$

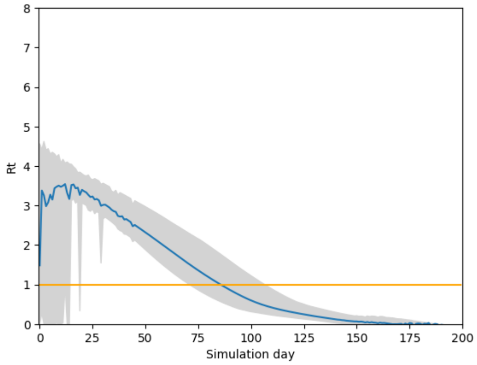


**(f)** $\alpha_i = 0.2$

**Fig S27.** Smoothed effective $R_t$ per day when varying heterogeneity in infectiousness, for the scenario without interventions. The blue line indicates the mean $R_t$ per day, while the gray area represents the interval in which 95% of observations lie.

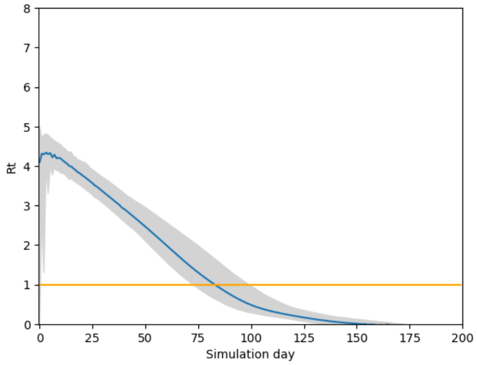


**(a)** Baseline

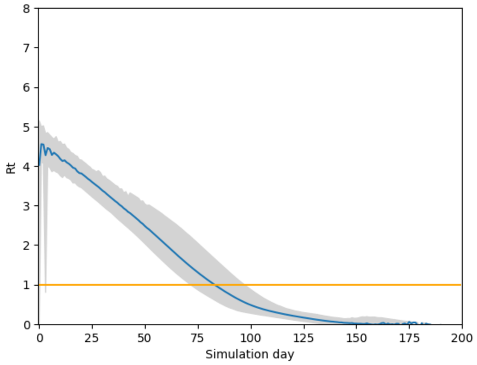


**(b)** $\alpha_c = 10$

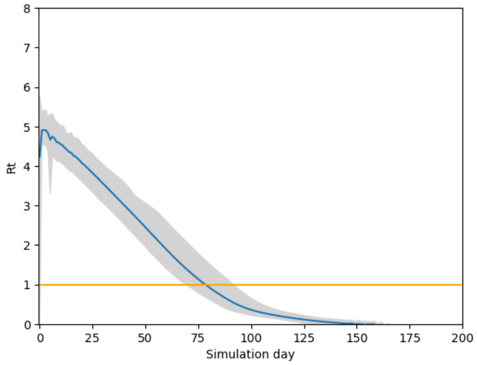


**(c)** $\alpha_c = 1$

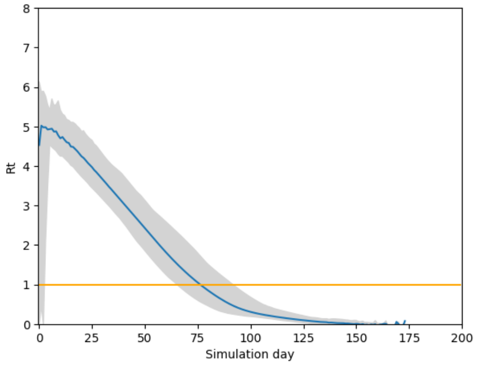


**(d)** $\alpha_c = 0.6$

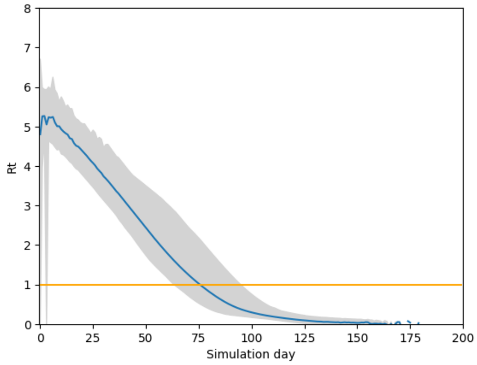


**(e)** $\alpha_c = 0.4$

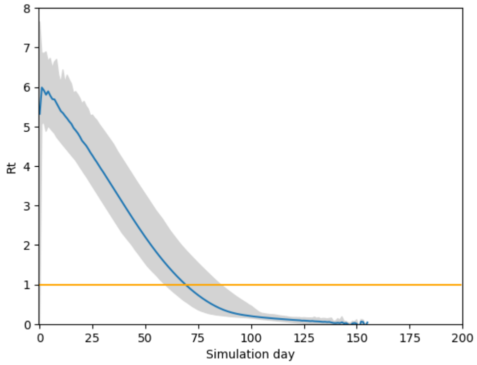


**(f)** $\alpha_c = 0.2$

**Fig S28.** Smoothed effective $R_t$ per day when varying heterogeneity in contact behavior, for the scenario without interventions. The blue line indicates the mean $R_t$ per day, while the gray area represents the interval in which 95% of observations lie.

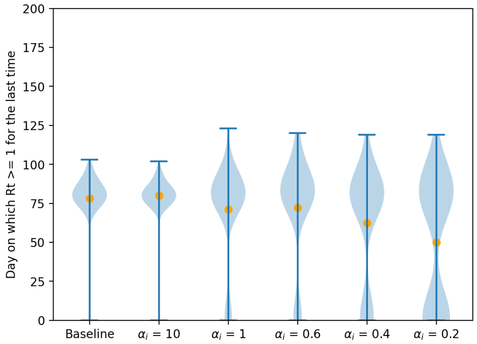


**(a)** Varying $\alpha_i$ for the Truncated Gamma distribution considered for the individual transmission probability.

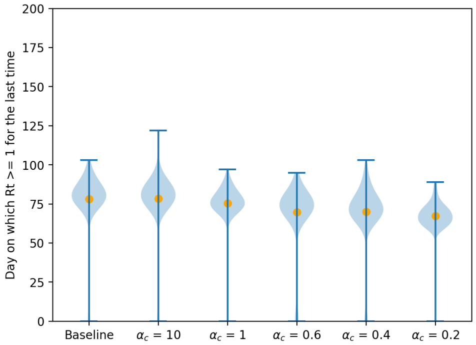


**(b)** Varying $\alpha_c$ for the Gamma distribution considered for the individual contact factor.

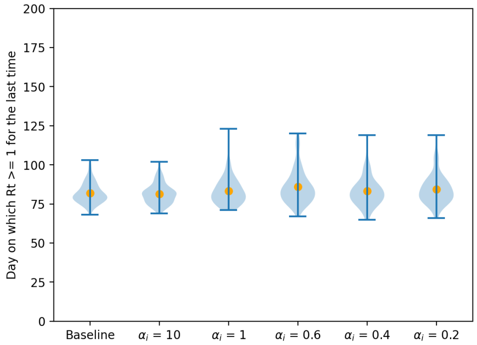


**(c)** Varying $\alpha_i$ for the Truncated Gamma distribution considered for the individual transmission probability. Runs with led to extinction ($< 20$ cases) were excluded.

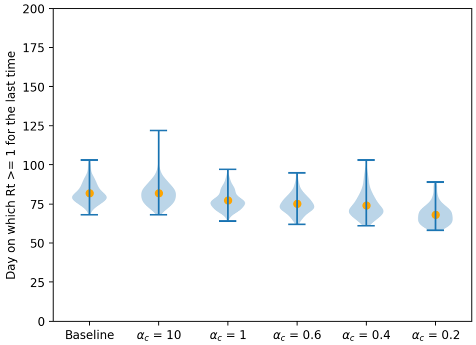


**(d)** Varying $\alpha_c$ for the Gamma distribution considered for the individual contact factor. Runs with led to extinction ($< 20$ cases) were excluded.

**Fig S29.** Violin plots for the day on which the herd immunity threshold is reached for the different scenarios without interventions, over all simulations runs (subfigure a–b) and only for simulations runs that generate more than 20 cases (subfigure c–d). Orange dots represent the means of the simulated values.

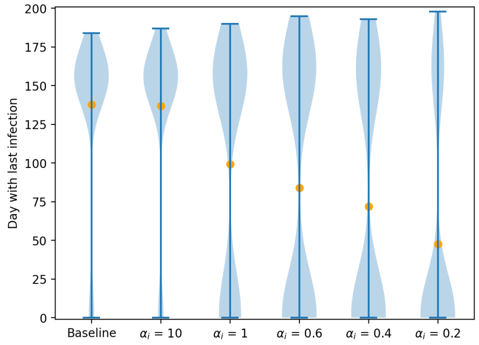


**(a)** Varying $\alpha_i$ for the Truncated Gamma distribution considered for the individual transmission probability.

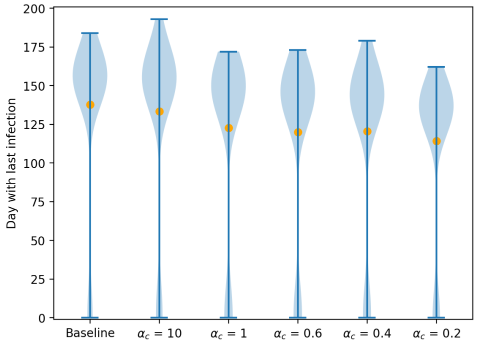


**(b)** Varying $\alpha_c$ for the Gamma distribution considered for the individual contact factor.

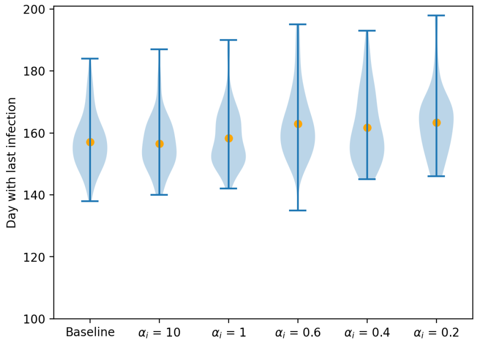


**(c)** Varying $\alpha_i$ for the Truncated Gamma distribution considered for the individual transmission probability. Runs with led to extinction ($< 20$ cases) were excluded.

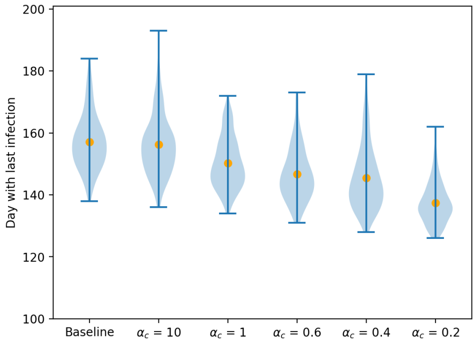


**(d)** Varying $\alpha_c$ for the Gamma distribution considered for the individual contact factor. Runs with led to extinction ($< 20$ cases) were excluded.

**Fig S30.** Violin plots for the day on which the last transmission event is observed for the different scenarios without interventions, over all simulations runs (subfigure a–b) and only for simulations runs that generate more than 20 cases (subfigure c–d). Orange dots represent the means of the simulated values.

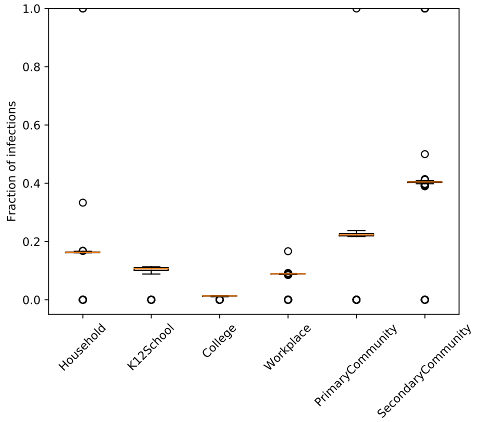


**(a)** Baseline

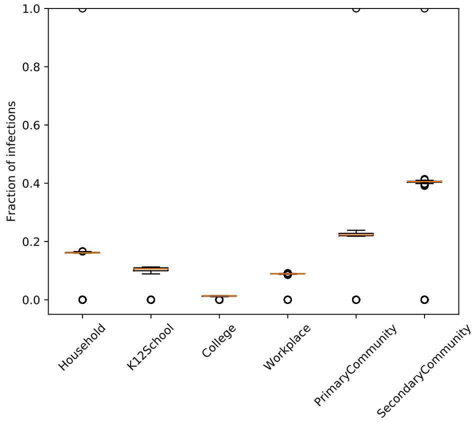


**(b)** $\alpha_i = 10$

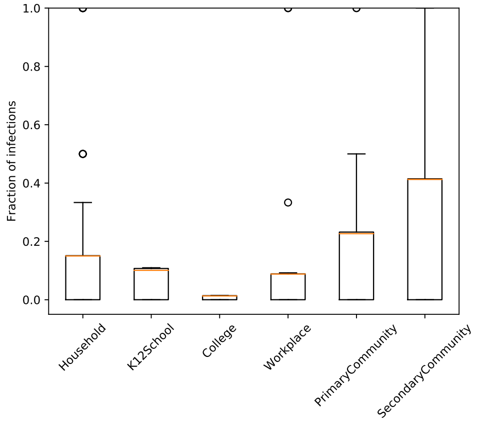


**(c)** $\alpha_i = 1$

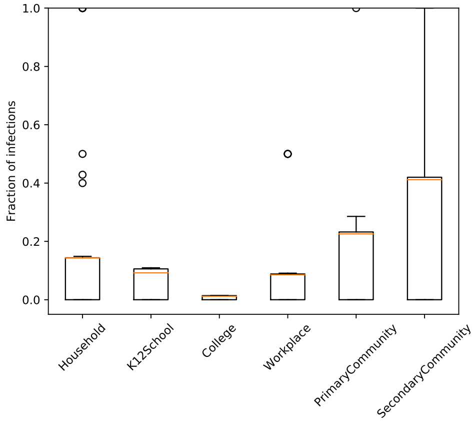


**(d)** $\alpha_i = 0.6$

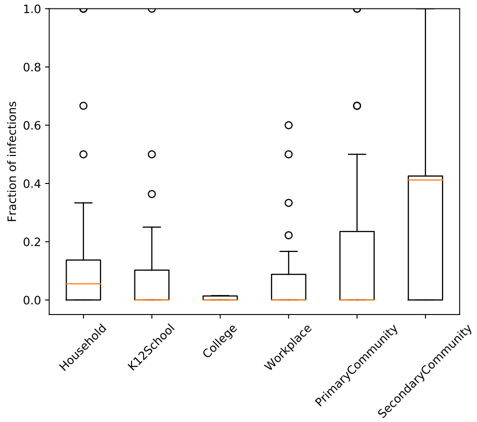


**(e)** $\alpha_i = 0.4$

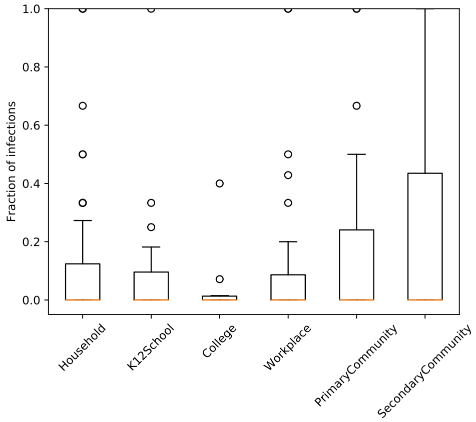


**(f)** $\alpha_i = 0.2$

**Fig S31.** Proportion of transmissions per location type for different values of $\alpha_i$ for the Truncated Gamma distribution considered for the individual transmission probability, for the scenario without interventions.

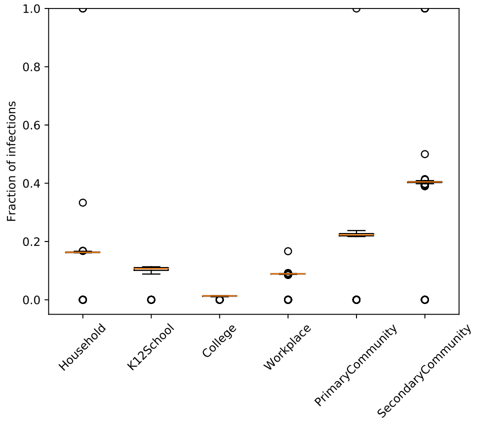


**(a)** Baseline

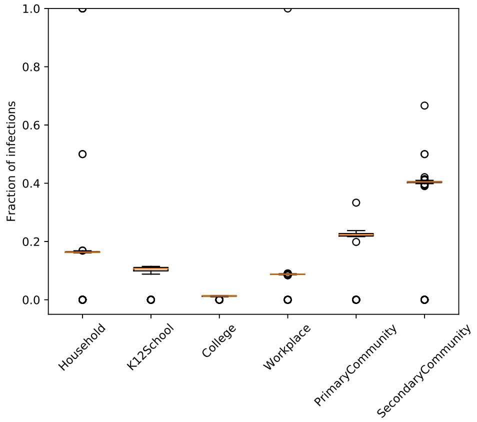


**(b)** $\alpha_c = 10$

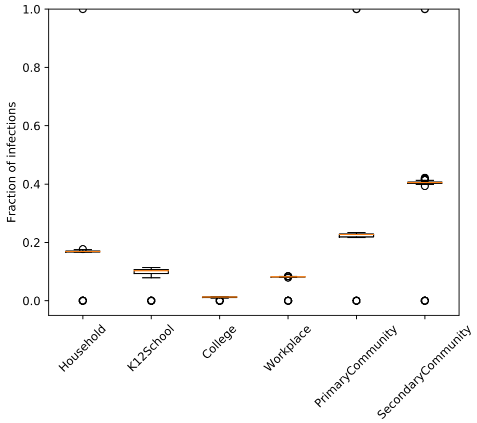


**(c)** $\alpha_c = 1$

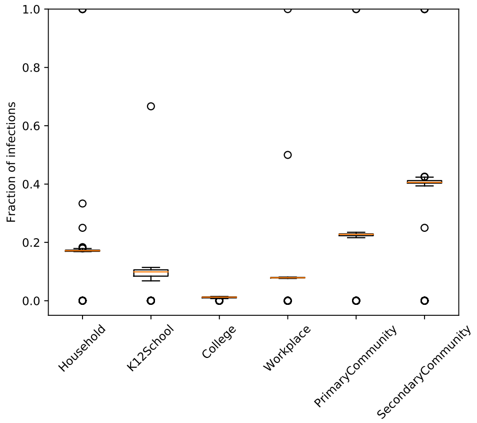


**(d)** $\alpha_c = 0.6$

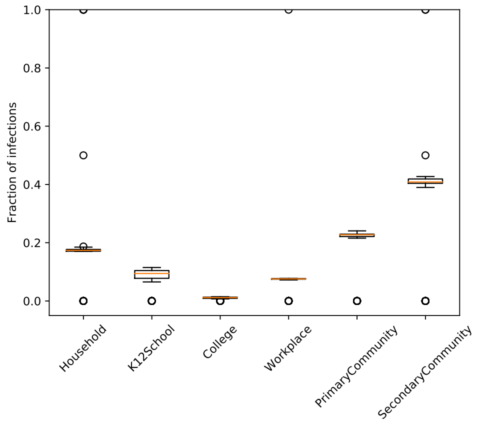


**(e)** $\alpha_c = 0.4$

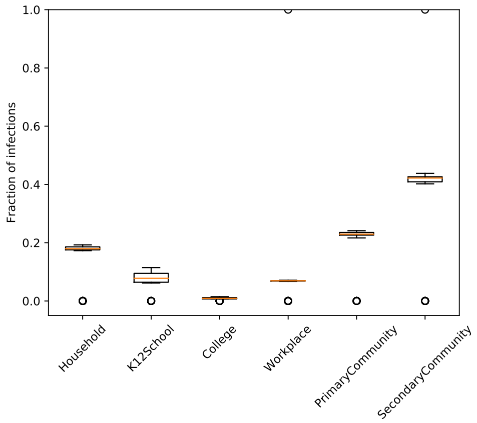


**(f)** $\alpha_c = 0.2$

**Fig S32.** Proportion of transmissions per location type for different values of $\alpha_c$ for the Gamma distribution considered for the individual contact factor, for the scenario without interventions.

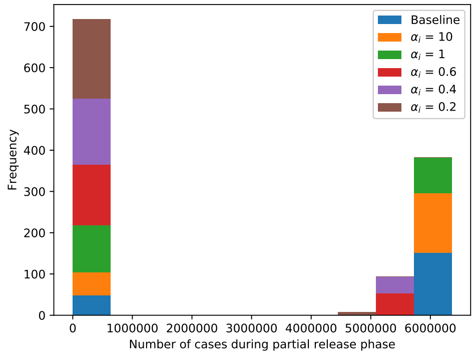


**(a)** Varying $\alpha_i$ for the Truncated Gamma distribution considered for the individual transmission probability.

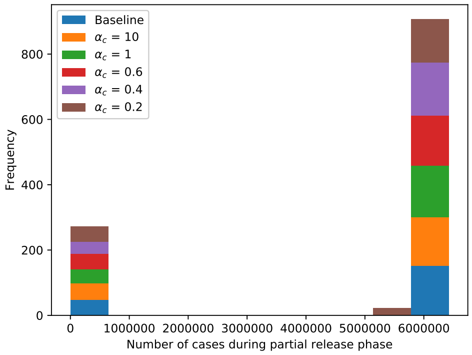


**(b)** Varying $\alpha_c$ for the Gamma distribution considered for the individual contact factor.

**Fig S33.** Histograms of the number of cases during the partial release phase for the different scenarios regarding infectiousness-related and contact-related heterogeneity, for the scenario with social distancing.

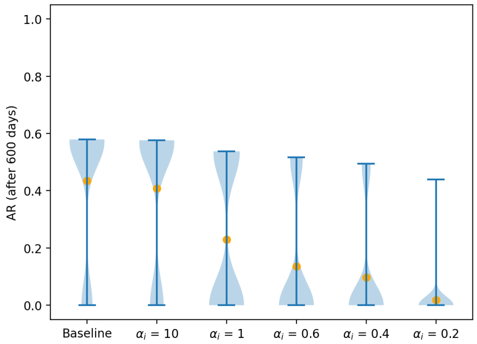


**(a)** Varying $\alpha_i$ for the Truncated Gamma distribution considered for the individual transmission probability.

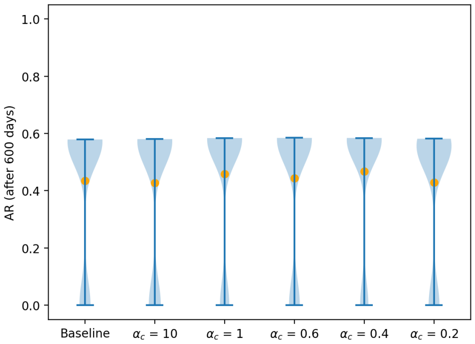


**(b)** Varying $\alpha_c$ for the Gamma distribution considered for the individual contact factor.

**Fig S34.** Violin plots for the attack rate over 600 days for scenarios investigating the infectiousness-related heterogeneity (panel a) and contact-related heterogeneity (panel b), with social distancing. The orange dots represent the mean attack rate across the simulation runs without extinction, i.e., simulation runs in which extinction occurs ($<$ 20 cases) were excluded.

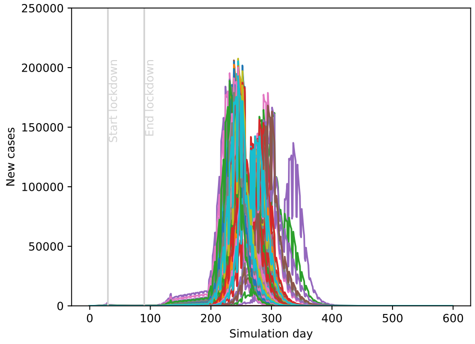


**(a)** Baseline

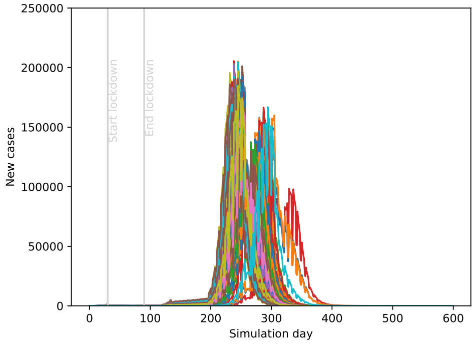


**(b)** $\alpha_i = 10$

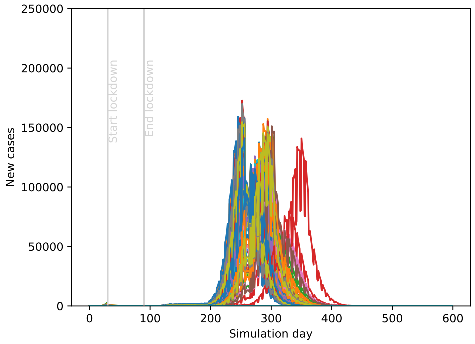


**(c)** $\alpha_i = 1$

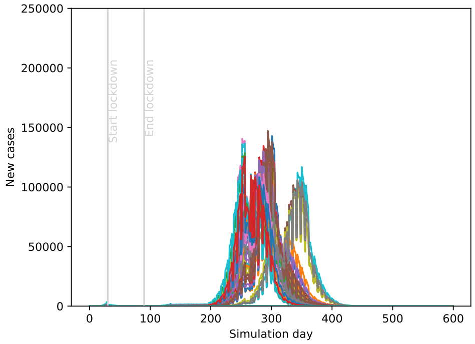


**(d)** $\alpha_i = 0.6$

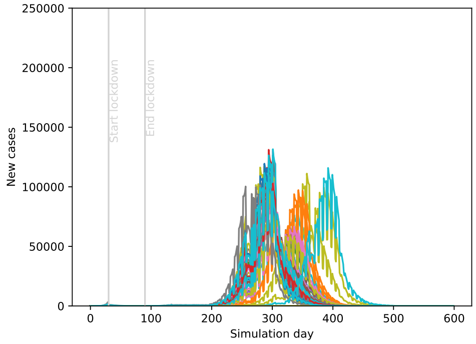


**(e)** $\alpha_i = 0.4$

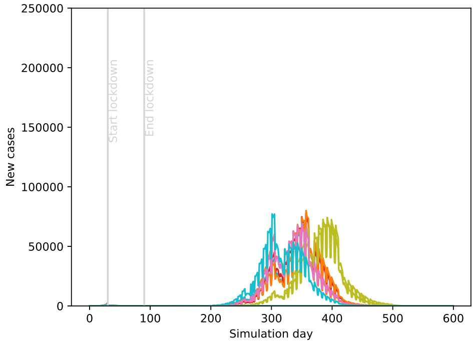


**(f)** $\alpha_i = 0.2$

**Fig S35.** Evolution of the number of new cases per day for different values of $\alpha_i$ for the Truncated Gamma distribution considered for the individual transmission probability, for the scenario with social distancing.

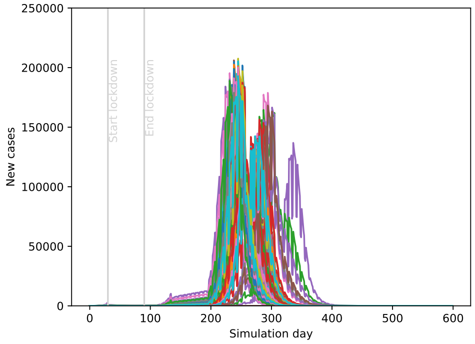


**(a)** Baseline

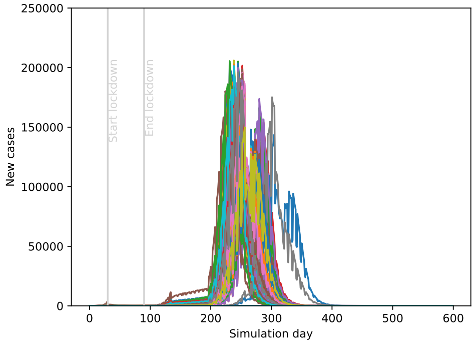


**(b)** $\alpha_c = 10$

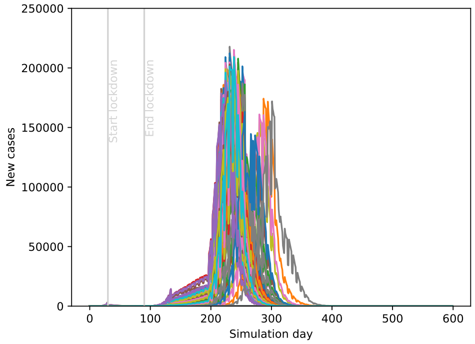


**(c)** $\alpha_c = 1$

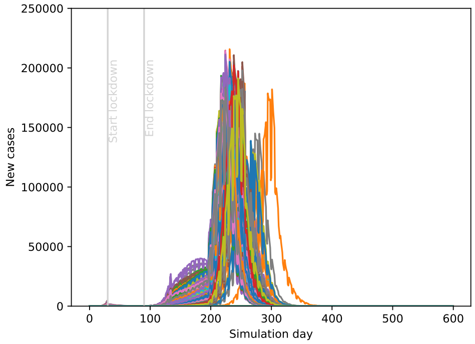


**(d)** $\alpha_c = 0.6$

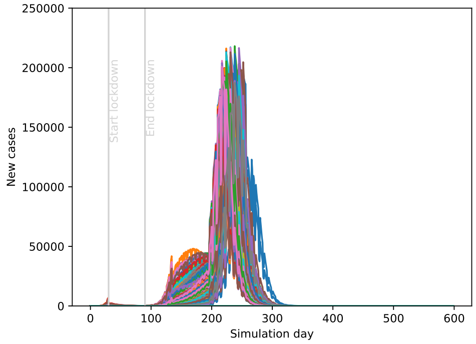


**(e)** $\alpha_c = 0.4$

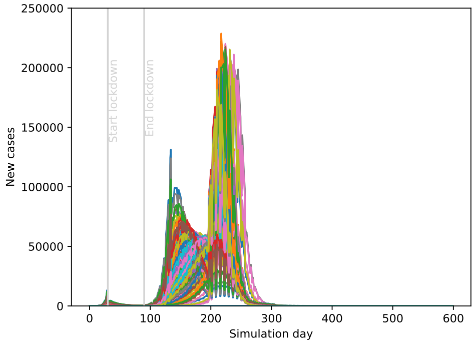


**(f)** $\alpha_c = 0.2$

**Fig S36.** Evolution of the number of new cases per day for different values of $\alpha_c$ for the Gamma distribution considered for the individual contact factor, for the scenario with social distancing.

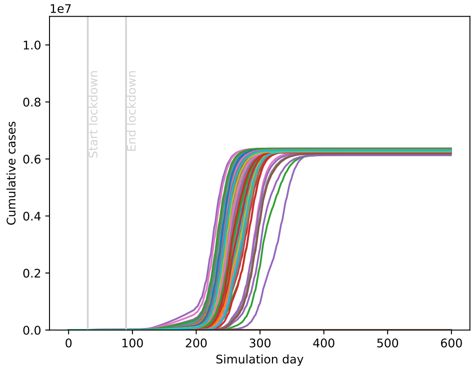


**(a)** Baseline

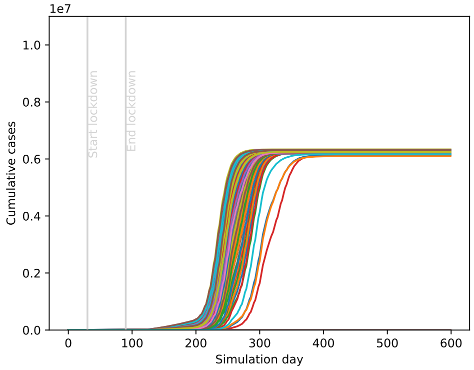


**(b)** $\alpha_i = 10$

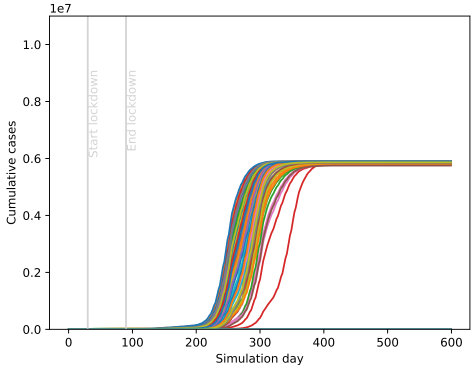


**(c)** $\alpha_i = 1$

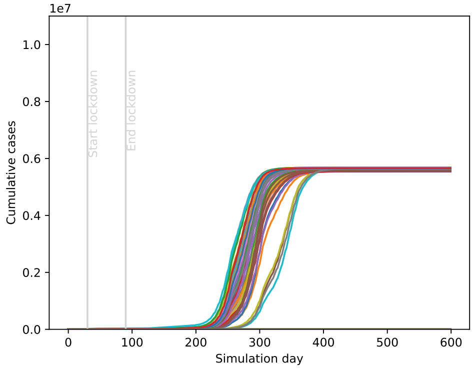


**(d)** $\alpha_i = 0.6$

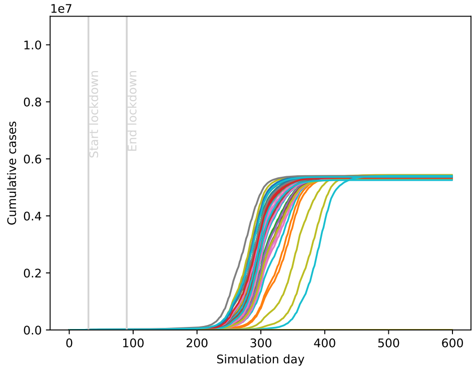


**(e)** $\alpha_i = 0.4$

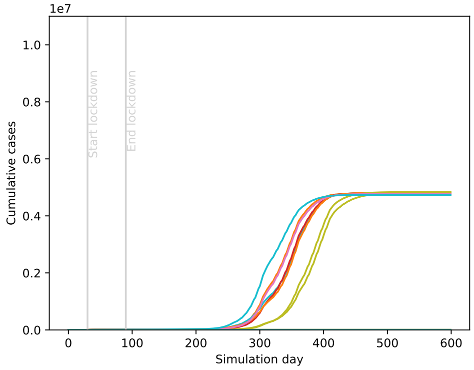


**(f)** $\alpha_i = 0.2$

**Fig S37.** Evolution of the cumulative number of cases per day for different values of $\alpha_i$ for the Truncated Gamma distribution considered for the individual transmission probability, for the scenario with social distancing.

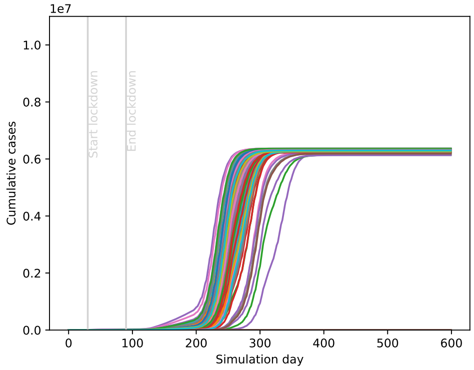


**(a)** Baseline

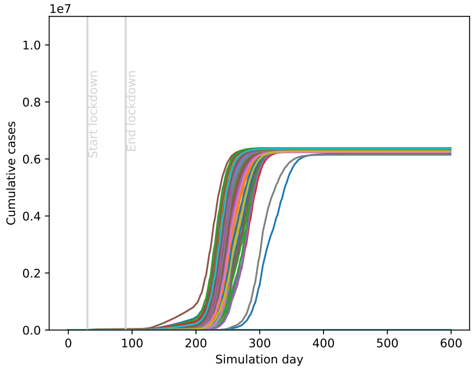


**(b)** $\alpha_c = 10$

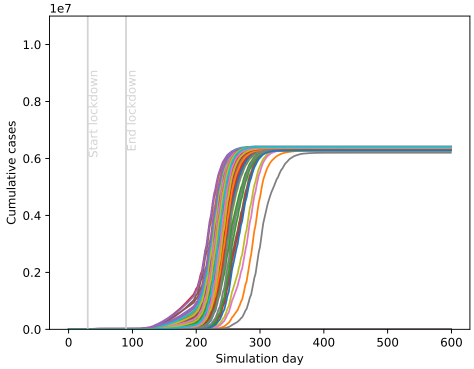


**(c)** $\alpha_c = 1$

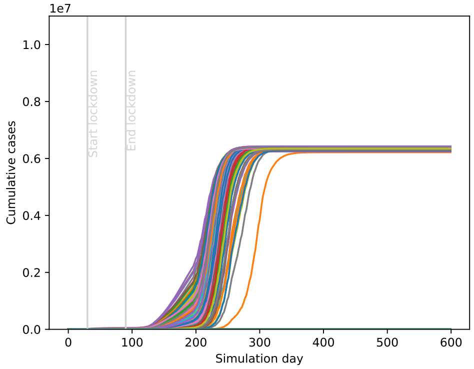


**(d)** $\alpha_c = 0.6$

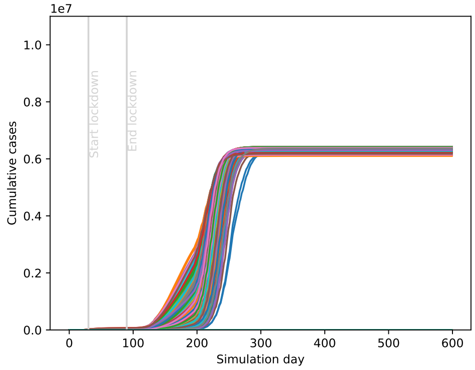


**(e)** $\alpha_c = 0.4$

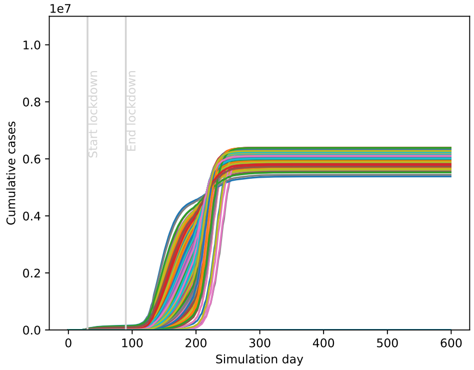


**(f)** $\alpha_c = 0.2$

**Fig S38.** Evolution of the cumulative number of cases per day for different values of $\alpha_c$ for the Gamma distribution considered for the individual contact factor, for the scenario with social distancing.

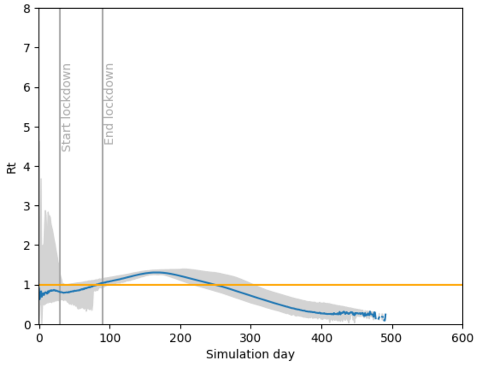


**(a)** Baseline

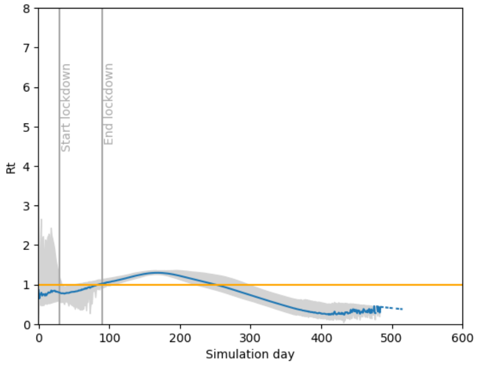


**(b)** $\alpha_i = 10$

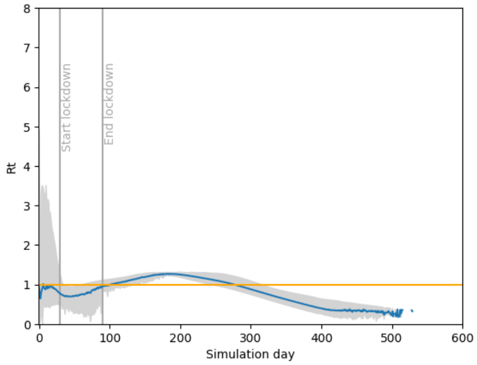


**(c)** $\alpha_i = 1$

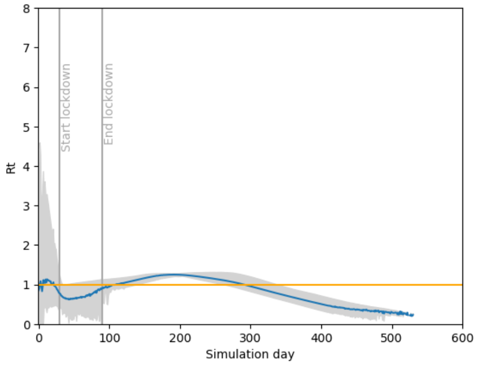


**(d)** $\alpha_i = 0.6$

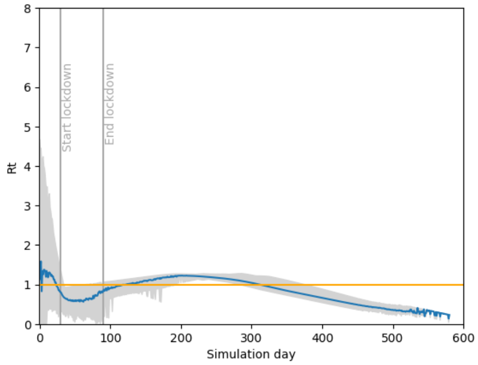


**(e)** $\alpha_i = 0.4$

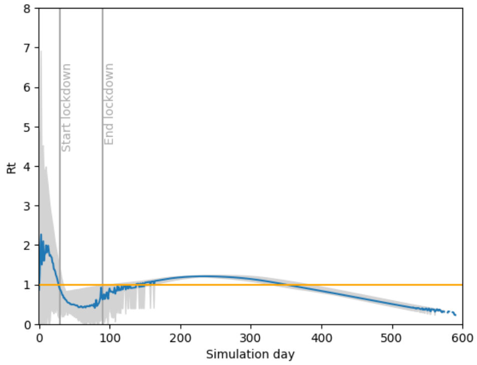


**(f)** $\alpha_i = 0.2$

**Fig S39.** Smoothed effective $R_t$ per day when varying heterogeneity in infectiousness, for the scenario with social distancing. The blue line indicates the mean $R_t$ per day, while the gray area represents the interval in which 95% of observations lie.

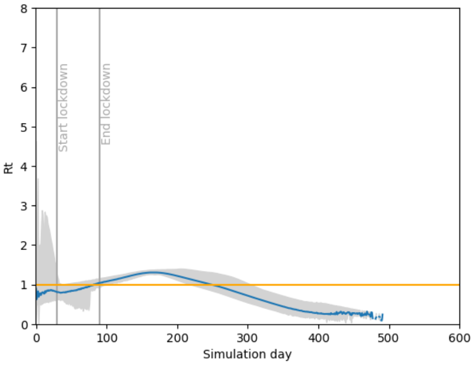


**(a)** Baseline

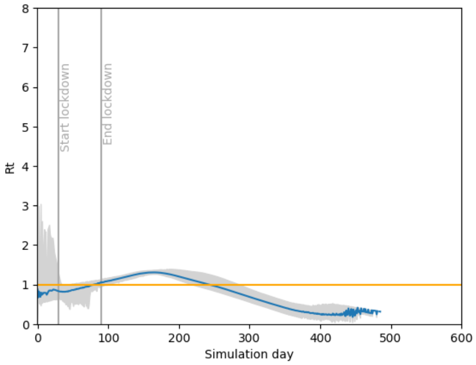


**(b)** $\alpha_c = 10$

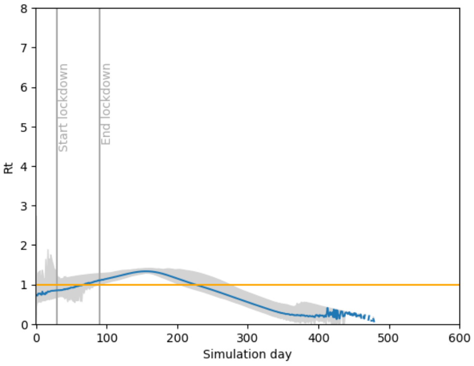


**(c)** $\alpha_c = 1$

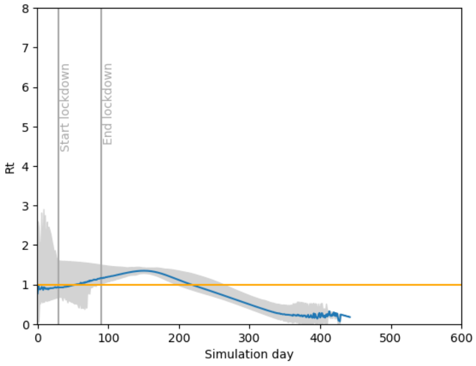


**(d)** $\alpha_c = 0.6$

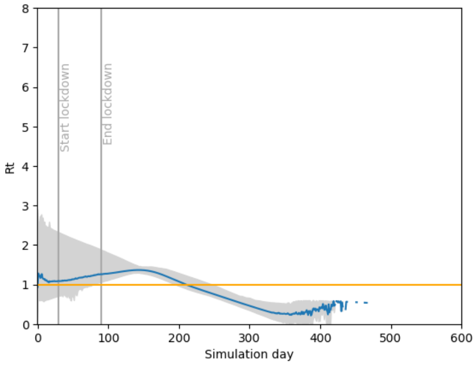


**(e)** $\alpha_c = 0.4$

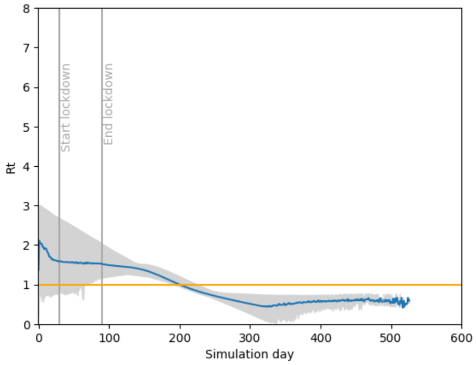


**(f)** $\alpha_c = 0.2$

**Fig S40.** Smoothed effective $R_t$ per day when varying heterogeneity in contact behavior, for the scenario with social distancing. The blue line indicates the mean $R_t$ per day, while the gray area represents the interval in which 95% of observations lie.

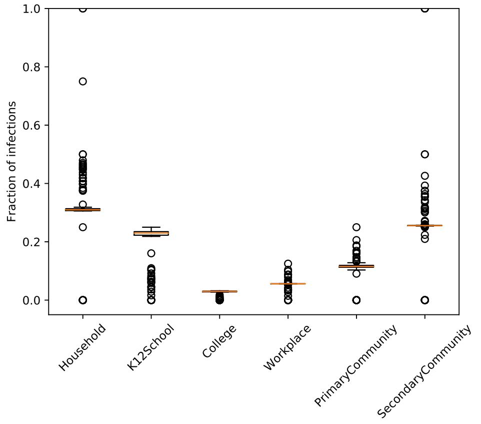


**(a)** Baseline

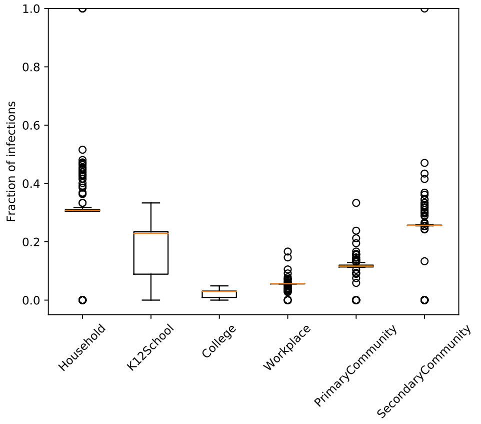


**(b)** $\alpha_i = 10$

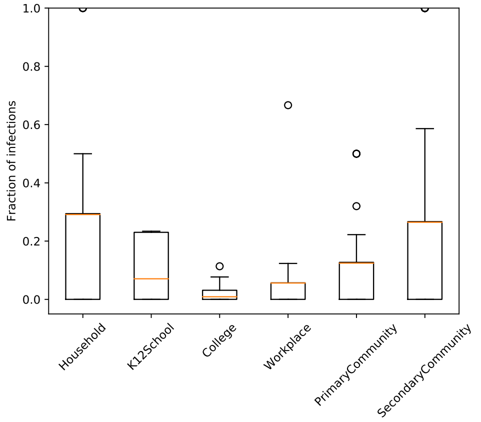


**(c)** $\alpha_i = 1$

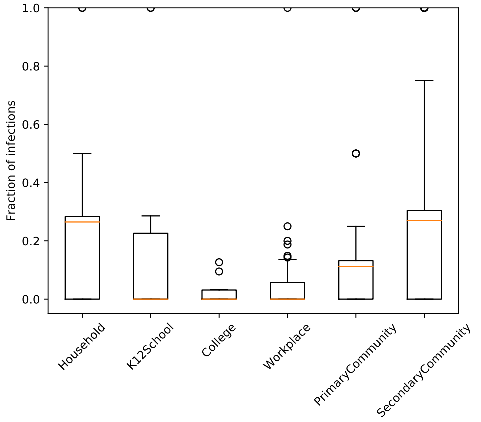


**(d)** $\alpha_i = 0.6$

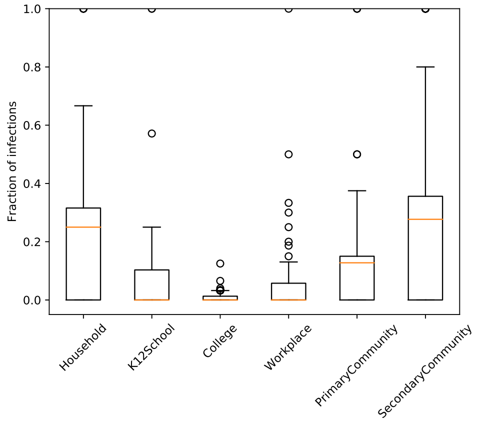


**(e)** $\alpha_i = 0.4$

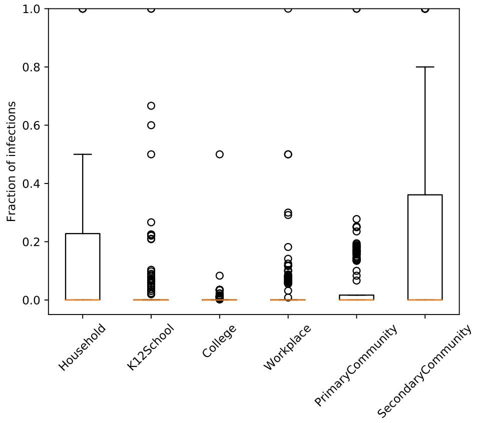


**(f)** $\alpha_i = 0.2$

**Fig S41.** Proportion of transmissions per location type for different values of $\alpha_i$ for the Truncated Gamma distribution considered for the individual transmission probability, for the scenario with social distancing.

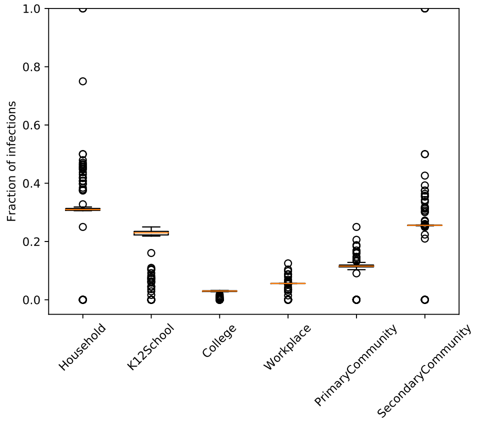


**(a)** Baseline

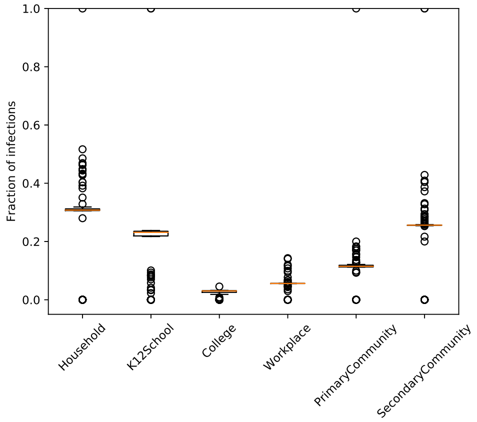


**(b)** $\alpha_c = 10$

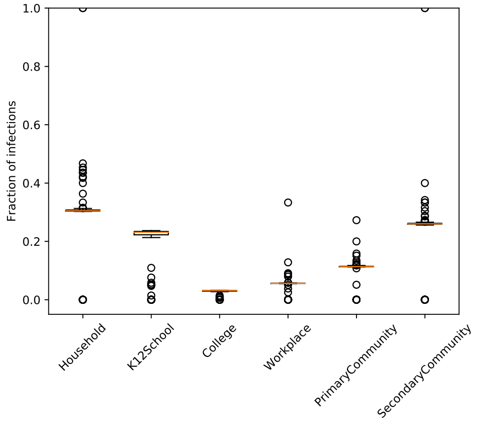


**(c)** $\alpha_c = 1$

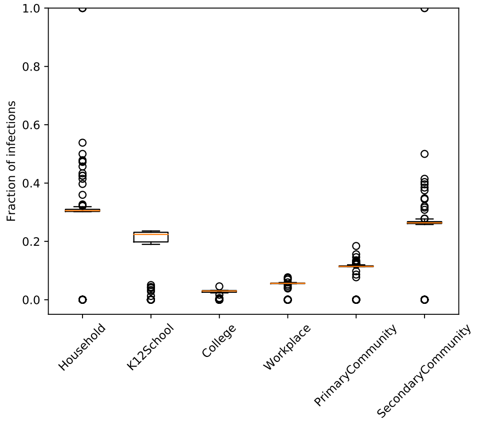


**(d)** $\alpha_c = 0.6$

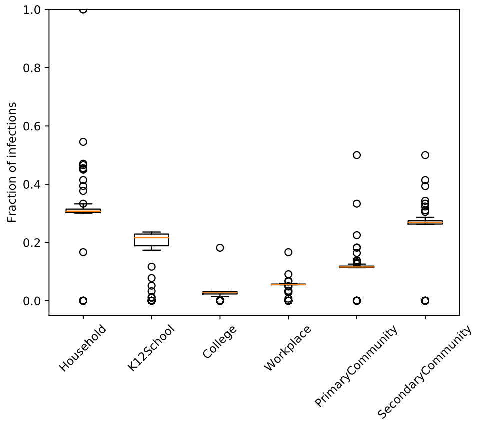


**(e)** $\alpha_c = 0.4$

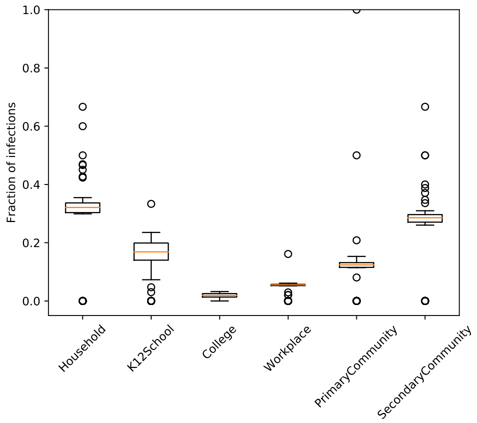


**(f)** $\alpha_c = 0.2$

**Fig S42.** Proportion of transmissions per location type for different values of $\alpha_c$ for the Gamma distribution considered for the individual contact factor, for the scenario with social distancing.

11. Lourenco J, Paton R, Ghafari M, Kraemer M, Thompson C, Simmonds P, et al. Fundamental principles of epidemic spread highlight the immediate need for large-scale serological surveys to assess the stage of the SARS-CoV-2 epidemic. MedRxiv. 2020;.

12. Wu JT, Leung K, Bushman M, Kishore N, Niehus R, de Salazar PM, et al. Estimating clinical severity of COVID-19 from the transmission dynamics in Wuhan, China. Nature medicine. 2020;26(4):506–510.

13. Kuylen E, Stijven S, Broeckhove J, Willem L. Social contact patterns in an individual-based simulator for the transmission of infectious diseases (stride). Procedia Computer Science. 2017;108:2438–2442.

14. Kremer C, Torneri A, Boesmans S, Meuwissen H, Verdonschot S, Vanden Driessche K, et al. Quantifying superspreading for COVID-19 using Poisson mixture distributions. Scientific reports. 2021;11(1):1–11.
